# X-Admix: An Interpretable Multimodal Cross-Attention Framework for Integrating Genotype, Local Ancestry, and Social Drivers of Health in Admixed African American Populations

**DOI:** 10.64898/2026.09.03.26362179

**Authors:** Nahian Tahmin, Lokesh K. Chinthala, Tesfaye B. Mersha, Robert L. Davis, Anahita Khojandi

## Abstract

Disease risk in admixed human populations is shaped by interactions among genotype, locus-specific ancestry, and the social environment, but predictive frameworks rarely model these three modalities jointly. We introduce X-Admix, an interpretable multimodal framework integrating genotype, local ancestry, and social drivers of health through structured pairwise cross-attention streams, softmax-gated fusion, and a Random Forest classifier. Unlike concatenation-based fusion, these directed streams learn conditional representations in which genotype is contextualized by local ancestry and social drivers of health, and local ancestry by social drivers of health. Leave-one-stream-out ablation decomposes predictive performance and top cross-modal pair candidates. Applied to 240 African American children with severe asthma from the BIG dataset, X-Admix predicted inhaled-corticosteroid response with mean area under the receiver operating characteristic curve 0.771 ± 0.072 across 10-fold cross-validation, whereas ridge logistic baselines performed near chance. This performance pattern replicated in 666 African American adults from the All of Us dataset under matched inclusion criteria. On the BIG dataset, a two-tier consensus pipeline yielded 932 top cross-modal feature pairs whose stream dependencies decomposed into stream-independent (14.7%), single-stream-conditional (28.7%), multi-stream-dependent (33.5%), and all-stream-dependent (23.2%). Without the genotype-local-ancestry stream, performance is unchanged, yet the top cross-modal pairs identified change, indicating that a stream’s contribution to interpretation and to prediction are separable: a stream redundant for prediction can still define the candidate interactions carried forward for discovery. To our knowledge, X-Admix is the first framework to jointly model the three data modalities via cross-attention in admixed cohorts, yielding an interpretable catalog of candidate interactions underlying inhaled-corticosteroid non-response.

**Author Summary:** Children and adults with asthma who share the same diagnosis—and even the same genetic variants -often respond very differently to inhaled steroid medications. In people of mixed ancestry, this variation reflects at least three things acting together: the genetic variants a person carries, the ancestral origin of the surrounding stretch of their genome, and the social and environmental conditions in which they live. Most predictive models treat these factors separately or simply add them together, which can hide how one factor changes the meaning of another. We built a framework, X-Admix, that instead lets each factor provide context for the others, so that a genetic variant can carry different information depending on the ancestry of its surrounding genomic region and a person’s environment. In African American children, and again in an independent group of African American adults, we found that X-Admix identified who would not respond to inhaled steroids more accurately than standard models built from the same information. We also obtained a ranked list of gene-ancestry-environment relationships, for example, air-pollution exposure acting together with immune-related genes-that suggest why treatment response varies and that can be tested directly in future studies.

## 1 Introduction

Multimodal modeling has become an increasingly popular strategy in biomedical research because clinically relevant phenotypes often emerge from the combined influence of heterogeneous factors rather than from any single data source [1–4]. This need is especially critical for admixed populations, where disease risk is shaped by interactions across multiple modalities: genotype captures allelic variation, local ancestry (LA) characterizes locus-specific ancestral context, and social drivers of health (SDOH) capture the environmental and structural factors that shape exposure and health outcome. Risk-relevant signal is therefore distributed across all three layers simultaneously, making modeling in admixed populations both challenging and still insufficiently addressed.

In African American (AA) admixed cohorts, conventional genomic analyses frequently underrepresent the full structure of disease risk. They rely on genotype alone or treat ancestry as a global covariate, overlooking the locus-specific inheritance that LA captures. Thus, two individuals may share the same genotype at a locus and similar global ancestry while carrying that genotype on genomic segments of different ancestral origin. Prior work demonstrates that genotype and ancestry signals are complementary rather than redundant [5–8], and that modeling them jointly can improve power and alter discovery relative to standard GWAS frameworks [5, 9–11]. For instance, local-ancestry-adjusted GWAS surfaced a functional CYP2C19 variant in African Americans that standard GWAS missed [12]. However, most of this literature focuses on locus-level association testing rather than predictive modeling, and rarely asks how genotype and LA can be integrated within a machine learning framework. This gap is especially significant in AA populations, where recent admixture creates heterogeneous ancestry blocks and where SDOH contribute additional risk not captured by genetics alone [13–15].

A central methodological challenge is that genotype, LA, and SDOH are heterogeneous data types whose effects are often nonlinear, conditional, and cross-modal [16–18]. This structure cannot be represented by conventional fusion strategies without collapsing modality-specific information. Traditional association models [19], mixed linear models [20–22], and related classical machine learning approaches [23, 24] remain essential to control population structure and estimate main effects, but are generally not designed to learn interactions between modalities that operate on different scales and carry different biological or social meaning. Structured cross-modal fusion via cross-attention addresses this by allowing one modality to selectively query another at the feature level, preserving modality-specific structure while learning conditional dependencies [25, 26]. This is particularly relevant when one modality provides locus-specific or contextual information for another (e.g., LA provides ancestral context, or SDOH provides exposure context) rather than simply co-varying with it [27–30]. This architecture is especially relevant for admixed populations, where genotype, LA, and SDOH may carry conditional, cross-modal structure that additive models represent only implicitly.

Modeling all three factors together also clarifies that ancestry and SDOH, though correlated, are not interchangeable. In admixed cohorts, shared ancestry can coincide with shared social exposures through structural racism, residential segregation, environmental inequality, and unequal access to care [31–33]. When SDOH are omitted, ancestry may inadvertently act as a proxy for these social exposures, conflating biological and structural pathways [34–36]. Explicitly including SDOH is therefore helpful to disentangle these contributions and represent disease risk more faithfully [14, 15, 37, 38].

Motivated by these requirements, we developed the X-Admix framework, an interpretable multimodal framework that integrates genotype, LA, and SDOH through structured cross-modal fusion. Unlike simple multimodal inclusion through concatenation, X-Admix uses three directed cross-attention streams to learn conditional representations in which genotype is contextualized by LA and SDOH, and LA is contextualized by SDOH. These separable streams and the explicit latent space support ablation- and attribution-based interpretation. We evaluated X-Admix for predicting response to inhaled corticosteroids (ICS) in a cohort of AA pediatric patients with severe asthma. Notably, AA children bear a disproportionate severe asthma burden and poorer ICS response than other populations [39–41]. These disparities underscore the clinical importance of developing ancestry-aware predictive models tailored to this high-risk population.

Three established comparators benchmark the multimodal framework in our setting. Regularized logistic regression (LR), specifically the *L*_2_-penalised (ridge) formulation, is a frequently used high-dimensional linear baseline in genomic prediction and admixed-ancestry risk studies [23, 24, 42]. Pairwise interaction testing via logistic models with explicit product terms is the established approach for detecting gene-environment and epistatic interactions [16, 43]. We combine these two specifications into three ridge-logistic baselines: main effects only, pairwise interactions only, and their union.

We further train and evaluate X-Admix on the All of Us (AoU) Research Program [44, 45]. AoU is the largest available admixed-population cohort with linked genomic and SDOH data [44–46]. Although this dataset consists exclusively of adults and is not a direct analogue of pediatric ICS pharmacology, it preserves the underlying cross-modal structure that X-Admix is designed to capture.

Additionally, the methodological design of X-Admix enables leave-one-stream-out ablation, in which each pairwise cross-attention stream is disabled to isolate its contribution [47, 48]. We apply this analysis to two separate outputs: predictive performance and the top cross-modal interactions, the feature pairs the model co-embeds, identified for interpretation. Removing a stream may not affect prediction performance yet shape which top candidate interactions are identified for downstream discovery [49, 50]. We therefore report stream-level dependencies alongside aggregate predictive performance.

This study makes three contributions: (*i*) to our knowledge, this is the first predictive framework to jointly model genotype, LA, and SDOH via cross-attention in an admixed cohort; (*ii*) it introduces a rigorous evaluation and interpretation approach that combines cross-validation (CV) with systematic per-stream ablation analyses; and (*iii*) it demonstrates the utility of the framework in two complementary cohorts: a pediatric AA severe asthma case study and a broadly aligned AoU adult cohort.

### 1.1 Literature Review

Admixed populations carry genomic regions with specific locus inherited from different ancestral background, and standard genomic methods may leave substantial cohort variation unmodeled [5, 9]. Adding locus-specific ancestral context through LA is one approach that can help address this shortcoming, and prior work demonstrates that genotype and LA contribute partially non-overlapping information and that their joint use improves performance over either source alone [5, 8, 51]. These methods were developed for locus-level association testing rather than for learning a shared multimodal representation. Building on this foundation, our prior severe asthma study extended the joint genotype-LA framing to a Single Nucleotide Polymorphism (SNP)-level predictive setting, demonstrating that LA was as informative as genotype in an AA pediatric cohort and that their combination outperforms either modality individually [52]. A natural next step is to incorporate SDOH alongside genotype and LA, since each modality may carry conditional rather than additive signal, and risk in admixed cohorts is distributed across biological, ancestral, and environmental contributions [14, 53, 54]. Aggregate polygenic and social risk combine largely additively for prediction [55], with each modality entering as a separate additive term; X-Admix complements this by conditioning each modality’s representation on the others through cross-attention, resolving the feature-level cross-modal structure into interpretable candidate pairs. To our knowledge, no prior framework has jointly modeled all three modalities in an interpretable predictive architecture; X-Admix addresses this by learning a shared multimodal representation as the basis for prediction and downstream interpretation.

The classical benchmark for our approach are ridge regression [56, 57] and logistic regression [16, 58, 59] on features, on raw pairwise interaction terms of the features, and on their union. Compared against the multimodal framework, these two classical families differ in their representational capacity. Ridge regression operates on a single design matrix of standardized features and learns only additive effects. Interaction-testing logistic models extend this design matrix with a pre-specified set of pairwise products, but their coverage of the interaction space is fixed in advance and each product is treated as an independent term with no shared structure across interactions. Neither approach learns its own interaction basis, and neither preserves a separation between modalities once the design matrix is assembled. We benchmark our approach against these alternatives in our admixed cohort.

Multimodal fusion strategies differ in how modality structure is handled: concatenation collapses modality-specific structure before any interaction is learned, while late fusion and gating-based approaches such as FiLM conditioning preserve modality boundaries but impose fixed conditioning relationships rather than allowing one modality to flexibly query another [26, 29, 60, 61]. Cross-attention sits in a different part of this space, in that the conditioning structure between modalities is itself learned at the feature level. This property has been leveraged across a range of biomedical settings, including multi-omics integration [62], single-cell multi-omics [63], imaging-genetics association [64], and clinical prediction under heterogeneous or partially missing inputs [65]. Across these applications, cross-attention preserves modality-specific structure while learning inter-modal dependence explicitly. We adapt the directional pairwise-stream design from multimodal language and affective computing [66–68], constructing one cross-attention stream per pair of genotype, LA, and SDOH. This provides a cleaner basis for downstream ablation and attribution than fusion strategies that entangle modalities at the input layer.

Interpreting cross-attention models in biomedical settings requires more than inspecting attention weights, which are insufficient as standalone explanations [69–71]. Recent methodological work on interpreting deep biomedical models therefore advocates layered attribution strategies, combining feature-level, latent-space, and pathway-level interpretation to derive biologically actionable outputs [72, 73]. Our approach incorporates this following principles to structure interpretability stage: Grad × Activation for the fused latent space [74] and SHAP [75] on the downstream classifier together define three attribution channels which intersections are used to surface cross-modal pairs.

A related practical consideration in small-cohort settings is training stability. When sample size is insufficient to support end-to-end training of a deep classifier, two-stage frameworks in which neural network derived embeddings feed a classical classifier such as Random Forest (RF) or LR offer a principled compromise [76, 77]. This design leverages the representational capacity of deep learning while relying on a downstream classifier with better characterized behavior under limited data [76, 77]. Two-stage approaches have been applied in genomics [78], clinical risk prediction [79], and imaging-based classification [80], and represent an established strategy when cohort size is constrained.

## 2 Methods

### 2.1 Data Description

Data are drawn from the Biorepository and Integrative Genomics (BIG) Initiative [81], a multi-site registry across the University of Tennessee Health Science Center, Le Bonheur Children’s Hospital, and Regional One Hospital (IRB #22-09164-NHSR). Informed consent was obtained from all participants or their legal guardians according to the Declaration of Helsinki. Table 1 summarizes the cohort characteristics. The inclusion criteria are as follows: asthma diagnosis by ICD code (493.x or J45.x), age ≥6 years, inhaled corticosteroid (ICS) prescription, and availability of whole exome sequencing (WES) data. ICS non-response (cases) is defined as an emergency department visit or hospitalization for asthma exacerbation within 24 months of initial ICS prescription; responders (controls) had no such event during the follow-up period. To increase the control pool, we additionally include participants without an asthma ICD code, consistent with the restriction of ICS prescribing to severe pediatric asthma [52, 82, 83]. The consort diagram is presented in Supplementary Figure S1.

**Table 1:** Demographic and clinical characteristics of the BIG Cohort.

| Characteristics | BIG Cohort |  |  |
| --- | --- | --- | --- |
|  | Overall | Cases | Controls |
| Patients, n (%) | 240 (100) | 122 (51) | 118 (49) |
| Female, n (%) | 117 (49) | 61 (52) | 56 (48) |
| Age, median (IQR) | 16 (13–18) | 16 (14–18) | 15 (12–18) |

### 2.2 Input Data Modalities

Three data modalities are integrated as inputs to the model.

#### Genotype

WES is performed on an Illumina NovaSeq 6000 using paired-end reads aligned to GRCh38 with BWA-MEM, with duplicates marked by Picard and variants called via DeepVariant v0.10.0 with joint calling through GLnexus v1.2.6.

#### LA

Genotype data are phased with SHAPEIT software [84] and processed through RFMix v2 [8] software against AFR (GWD, YRI, MSL) and EUR (CEU, TSI) reference panels from the 1000 Genomes Project [85], yielding locus-specific ancestry assignments across each participant’s genome.

#### SDOH

We geocode SDOH variables by mapping each participant’s residential address to a Census tract and linking the corresponding area-level exposure to the participant record. This area-level encoding represents environmental and structural exposures that act in the residential neighborhood rather than directly on the individual. Census tract linkage provides finer spatial resolution than ZIP-code geocoding, reducing exposure misclassification within heterogeneous areas.

For genotype features, PLINK [86, 87] is used to filter SNPs by minor allele frequency (< 5%), Hardy-Weinberg equilibrium (*α* = 0.05), and LD pruning (50 kb window, step 5, *r*^2^ = 0.2), then ordinally encoded (1/2/3) with *∼*2% missing calls imputed by *k*-nearest neighbors (*k* = 5). For LA features, RFMix v2 [8] is used to code LA segments as African (1) versus non-African (0), averaged across haplotypes per segment, and assigned to each SNP in that segment. Because the BIG cohort is small relative to the number of candidate variants, we constrain the LA candidate space using biological priors, which stabilizes feature selection in this setting [88, 89]. Candidate variants for the LA modality are therefore first restricted to a curated set of neuroimmune and asthma-pathway genes, assembled by querying curated pathway annotations from the Gene Ontology Biological Process [90, 91], KEGG [92], and Reactome [93] databases, accessed as gene-set libraries through Enrichr [94, 95], and retaining genes from pathways matching predefined asthma and neuroimmune term lists. Candidate SDOH variables are drawn from prior literature on social drivers of asthma severity [96–100].

Subsequently, feature selection is performed independently for each modality within each outer CV fold to prevent information leakage. The procedure combines biological prior knowledge with a two-pipeline stacked-ensemble strategy adapted from our prior work [52] and is summarized in Figure 1. For each modality, candidates are passed to a two-pipeline ensemble (Figure 1): an *L*_1_-regularized LR pipeline and an RF pipeline [52]. Each pipeline ranks features by a composite score combining permutation-based importance and mean absolute SHAP attribution within each fold.

**Figure 1:**
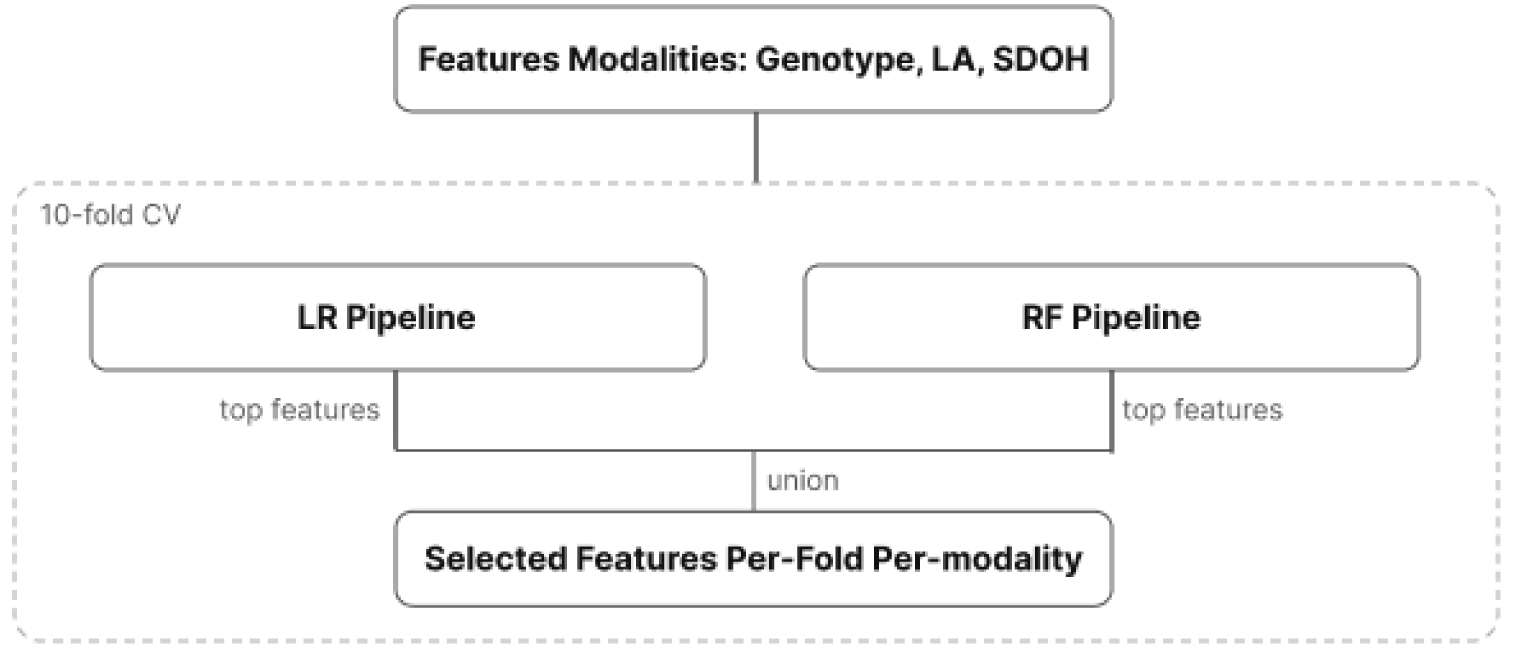
Per-fold feature selection. Within each outer CV fold, candidates from each modality (genotype, LA, SDOH) are passed through two complementary pipelines: an LR pipeline using *L*_1_-regularized (Lasso) logistic regression and an RF pipeline using a Random Forest classifier with modality-appropriate feature pre-screening. Both pipelines run under inner 10-fold CV for hyperparameter and threshold tuning, and the per-modality shortlist is defined as the union of the top features retained by the two pipelines. Adapted from [52].

For genotype, the modality-specific shortlist is defined as the union of features retained by the LR and RF pipelines, corresponding to approximately 15% of the *∼*95 selected variants per fold. For LA, all candidates, approximately 240 variants per fold, are passed to the cross-attention model without further filtering. We adopt this approach because LA signal is distributed across many variants, so that any individual variant carries small importance and SHAP magnitudes, making magnitude-based selection uninformative. For SDOH, this procedure narrow the candidate set to four variables: PM2.5 exposure, Social Vulnerability Index (SVI), Area Deprivation Index (ADI) national rank, and urban/rural status; categorical SDOH variables are label-encoded and numeric SDOH variables entered the model on their original scale. The resulting per-modality feature sets are passed as input to the cross-attention pipeline, and per-fold feature pool sizes are reported in Supplementary Table S1.

#### Overview of the X-Admix Framework

We develop a 3-stage multimodal framework called X-Admix to predict ICS non-response from genotype, LA, and SDOH data (Figure 2). In Stage 1, a neural fusion architecture learns a shared latent representation by computing pairwise cross-attention across the three modalities. In Stage 2, this latent representation serves as input to a downstream classifier for final risk prediction. Finally, Stage 3 is post-hoc interpretability analysis accumulated across multiple steps. All analyses are conducted under 10-fold stratified CV with predefined fold assignments [52] matching the feature selection folds. The performance metrics, Receiver Operating Characteristic curve (AUC), precision, recall, are computed on the test-fold partition, and we report fold-wise means *±* standard deviations (SD).

**Figure 2:**
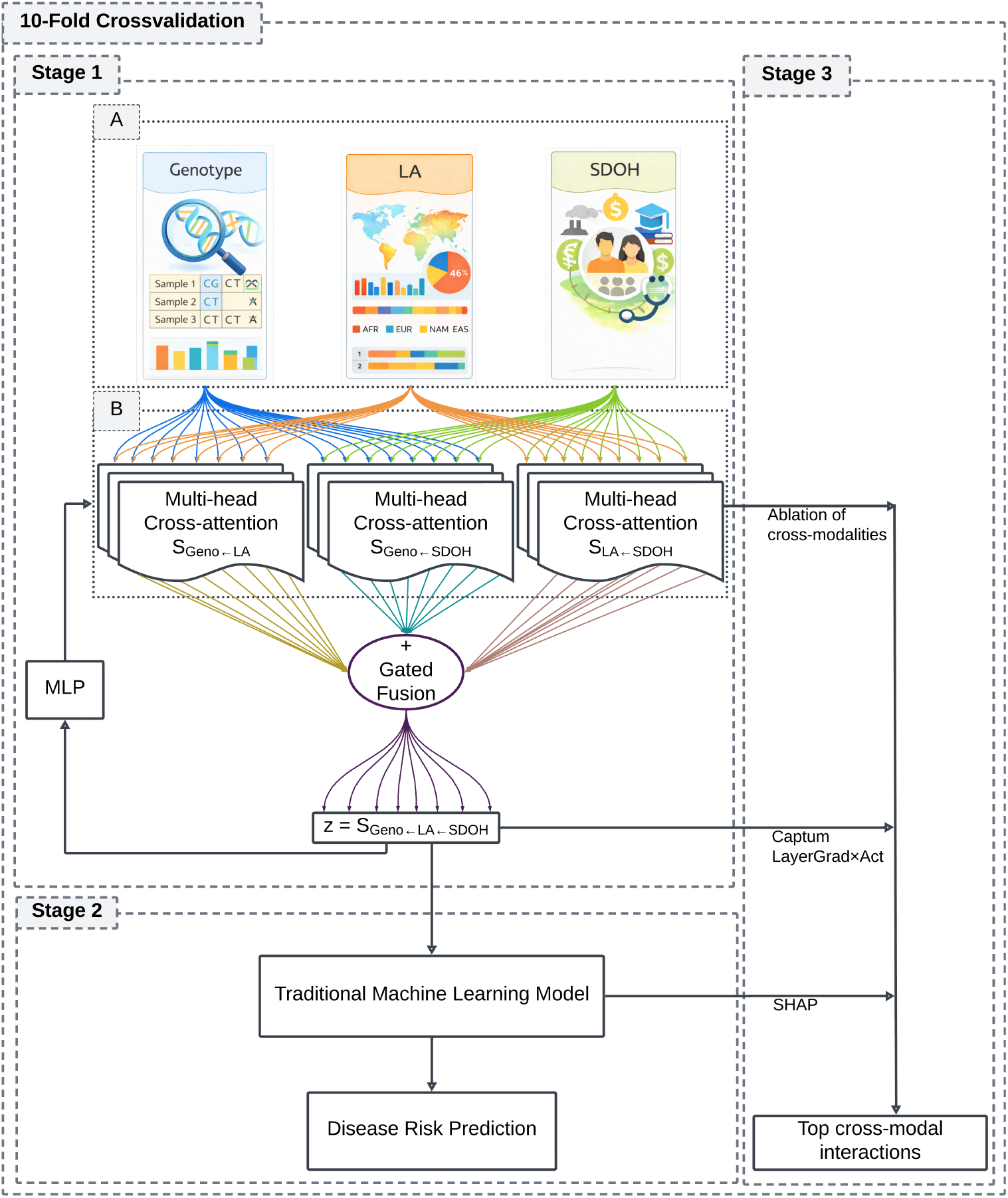
The *X-Admix* framework for cross-modal integration of genotype, LA, and SDOH. In stage 1, three modality-specific token embeddings feed three pairwise multi-head cross-attention streams, *S_Geno←LA_*, *S_Geno←SDOH_*, and *S_LA←SDOH_*, combined through a learnable softmax-gated fusion layer to produce a shared latent representation *z*. An MLP head attached to *z* constrains the fused space toward predictive structure. In Stage 2, *z* is passed to RF for final risk prediction. Stage 3 applies interpretability techniques to jointly yield rankings of cross-modal interactions.

The fusion architecture consists of three pairwise cross-attention streams (Figure 2), each capturing a distinct mode of inter-modal interaction: *S_Geno←LA_*, in which genotype attends to ancestry context; *S_Geno←SDOH_*, in which genotype attends to SDOH exposures; and *S_LA←SDOH_*, in which LA attends to SDOH exposures. In general, *S_X←Y_* denotes the cross-attention stream in which modality *X* is the query and modality *Y* is the key/value. We refer to the complete pipeline incorporating all three streams as the *S_integrated_* pipeline, and two-stream ablations (*−S_Geno←LA_*, *−S_Geno←SDOH_*, *−S_LA←SDOH_*) retain the two unnamed streams and disable the named one. Each ablation is retrained from scratch under the same hyperparameter sweep and 10-fold cross-validation as *S_integrated_*. Accordingly, *S_integrated_* jointly combines three directional pairwise conditional representations rather than a single formal genotype × LA × SDOH interaction coefficient.

### 2.3 Stage 1: Fusion Architecture

#### Input Embedding

Each feature value is represented as a single scalar token and projected into a shared latent space of dimension *d*_model_, the working dimension of the cross-attention model, through modality-specific linear layers. Genotype and LA tokens additionally carry a tag describing where the variant lies in the genome. This lets the model distinguish two variants that happen to share the same value and sense how close together two variants sit. The chromosome label is converted to an integer identifier and mapped to a learned chromosome embedding, and the base-pair coordinate is encoded separately. Because raw coordinates span an enormous range, each position is first *z*-scored to give a standardized value *p̃*, which is then passed through a Fourier positional encoding analogous to those used in standard transformers [25, 101]. This yields a fixed-length encoding PE(*p̃*) in which nearby positions receive similar values and distant positions receive distinct ones. For example, the SNP chr12_26443685_C_T has *p̃ ≈ −*0.35 and yields the computed encoding PE(*p̃*) *≈* [*−*0.343*, −*0.644*, −*0.985*, −*0.335, 0.939, 0.765, 0.170*, −*0.942], whereas chr13_21155813_T_TA at *p̃ ≈* 0.78 yields a clearly different encoding PE(*p̃*) *≈* [0.703, 1.000, 0.022*, −*0.043, 0.711, 0.011*, −*1.000, 0.999]. The final genotype or LA token is the sum of three pieces projected to *d*_model_: the feature value embedding, the chromosome embedding, and the positional encoding PE(*p̃*).

SDOH features receive fixed identity encoding derived deterministically from variable names, which keeps every SDOH variable distinguishable even though it is represented by a single value. For example, PM_2.5_ is assigned 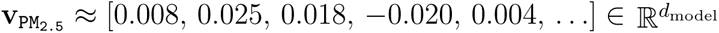, obtained by seeding a Gaussian draw from a 32-bit hash of the name. The final SDOH token is the element-wise sum of this identity encoding and the embedding of the feature value, giving one *d*_model_-dimensional token per feature.

#### Cross-Attention Streams

Each of the 3 pairwise streams (*S_Geno←LA_*, *S_Geno←SDOH_*, *S_LA←SDOH_*) is implemented as a standard multi-head cross-attention layer. Each stream involves two stages of multi-head attention. In the first stage, the query modality attends to the key/value modality through standard multi-head cross-attention, producing a sequence of attended representations (one per query token, each now informed by the other modality). In the second stage, a learnable classification (CLS) token queries this attended sequence through a dedicated multi-head attention layer, compressing the variable-length sequence into a single *d*_model_-dimensional summary vector per stream. This pooling step is necessary because the gated fusion layer requires fixed-size inputs from each stream, and attention-based pooling allows the model to learn which parts of the attended sequence are most relevant to aggregate, as opposed to a static mean pooling. We applied strict dropout to the pooled vectors before fusion to regularize the cross-modal summaries independently of the attention modules.

#### Gated Fusion

The 3 pooled stream vectors are combined via a learnable softmax gate. The gate produces weights *w*_1_*, w*_2_*, w*_3_ (with Σ*_i_w_i_* = 1) and forms both a weighted mixture (*w*_1_*s_Geno←LA_* + *w*_2_*s_Geno←SDOH_* + *w*_3_*s_LA←SDOH_*) and an unweighted sum (*s_Geno←LA_* + *s_Geno←SDOH_* + *s_LA←SDOH_*). These are concatenated, linearly projected to *d*_model_ dimensions, and passed through a supervised Multilayer Perceptron (MLP) to produce the fused latent representation *z*.

#### Learned Representation

During representation learning, the MLP classification head takes z as its input and maps it to the training logit. It consists of layer normalization, a linear bottleneck (width = max(*d*_model_*/r,* 32) where *r ∈ {*2, 4*}*), GELU activation, dropout, and a final linear output to a single logit. The bottleneck restricts the classification head’s capacity so that the model cannot memorize the prediction task in its final layers. This forces the preceding fusion layers to build more compact representation *z* that is predictive. The *z* is passed to a traditional machine learning classifier, which operates on the full *d*_model_-dimensions *z* without any bottleneck.

#### Encoder Training

The MLP is optimized with binary cross-entropy loss using AdamW with cosine-annealed learning rates, and its hyperparameters were tuned within a Bayesian sweep in Weights & Biases (W&B) [102]. Within each outer fold, training followed a two-step procedure that separates the choice of training duration from final fitting. A small stratified inner validation fold, sized to match the outer-test fold, was first used only for early stopping in training the MLP in fusion architecture. The MLP is then reinitialized and refit on the full outer-training fold for that fixed number of epochs; this refit model served as the fold’s final encoder, supplying the fused representation *z* used for the downstream prediction.

### 2.4 Stage 2: Traditional Machine Learning

The fused representation *z* is used as input to a RF classifier for the downstream prediction task. To avoid configurations favored by a single fortunate split during hyperparameter tuning, each candidate is scored by the RF’s mean inner-fold validation AUC across five repeated inner train/validation splits, penalized by its variability across those splits. We use an RF rather than the MLP head for final classification because the MLP head functions only as a training-time constraint that forces *z* to be linearly predictive, whereas a tree ensemble is better characterized than a parametric deep classifier in the limited-data setting and is more interpretable than the MLP. This two-stage design, in which a neural encoder supplies representations to a classical classifier, follows an established strategy for stabilizing prediction when cohort size constrains end-to-end training [76–78].

### 2.5 Stage 3: Interpretability Analyses

Interpretation proceeds through two complementary layers, each targeting a different level of the framework hierarchy (Figure 3). To identify which dimensions of the fused representation *z* are most influential for prediction, Captum LayerGradient × Activation [103] (Grad × Activation) is applied at the identity layer that exposed *z*, producing a per-sample attribution score for each *z*. In parallel, SHAP [75] is applied to the RF classifier to rank the *d*_model_ latent *z* by their contribution to the final prediction. For each CV fold, the top *N* = 30 fused *z* are filtered by the composite ranking by these methods.

**Figure 3:**
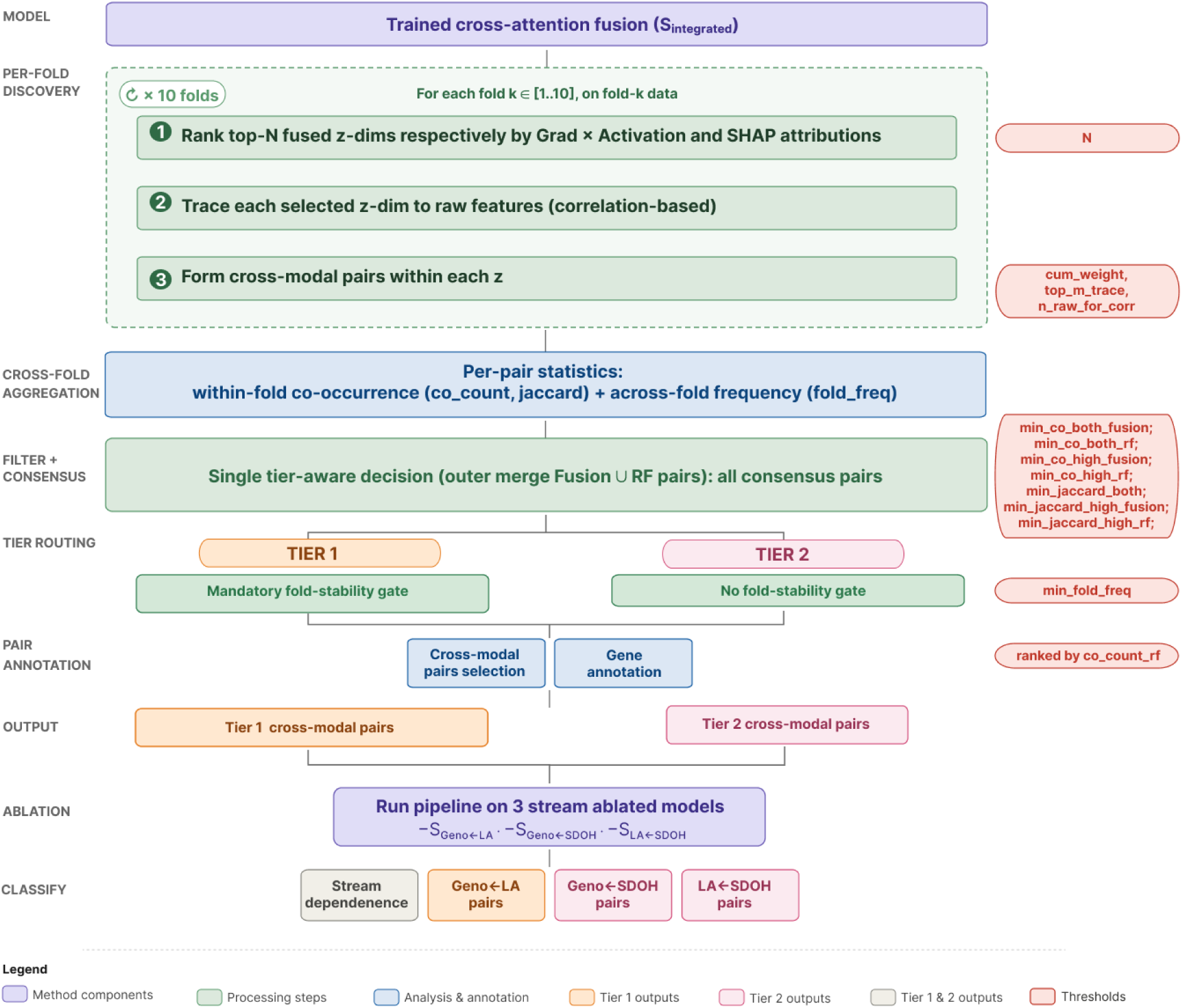
Interpretability structure for cross-modal pair discovery. The diagram traces the pipeline from per-fold discovery on the trained *S_integrated_* pipeline through cross-fold aggregation, per-method consensus, tier routing, pair annotation, and the final ablation classification step that assigns each pair to a stream-dependency category. The pipeline is run once on *S_integrated_* and once on each of the three single-stream ablations (*−S_Geno←LA_*, *−S_Geno←SDOH_*, *−S_LA←SDOH_*), allowing every consensus pair to be classified by its stream dependence for all ablations.

The heuristics at this stage are design choices rather than tuned quantities: retention parameters follow from the geometry and functionality of each channel’s candidate set. The same values are applied across folds, ablations, and both cohorts. Each retained *z_k_* is then traced to its raw features and corresponding modalities by correlation. Grad × Activation is computed through the MLP head and SHAP through the downstream RF, but both operate on the latent space of the same trained encoder, and both link a dimension to raw features by the absolute correlation between that dimension’s activation and the raw feature value. The candidate sets over which this correlation is applied, however, differ. Under Grad × Activation, the activation vector of *z_k_* on each fold’s test partition is correlated (Pearson) with the raw value of every genotype, LA, and SDOH feature, and the 15 features with the highest absolute correlation are retained per fold; these per-fold lists are pooled across folds, each feature is scored by its mean absolute correlation, and the 10 top-ranked features are carried forward. Correlation therefore selects the candidate set, and each dimension carries its own. Under SHAP, TreeExplainer is applied to the downstream RF over the latent dimensions; raw features receive no direct SHAP value and are instead ranked by a back-mapped score that sums *|*corr(*z_j_,* raw)*|* weighted by the mean *|*SHAP*|* of *z_j_* over all dimensions. A raw feature therefore ranks highly only if it tracks dimensions the classifier actually relies on. The 120 top-ranked raw features are retained as a single shared pool, independently of which dimensions are top-ranked, and each top-ranked *z_k_* is then correlated against this pool, producing carried forward sets for each dimension. Grad × Activation and SHAP thus serve to identify which latent dimensions are important for prediction, while this correlation step identifies the raw features associated with each such dimension. In both cases, within each dimension separately, features are retained in order of absolute correlation until their cumulative normalized correlation weight reaches 70% of that dimension’s carried-forward set, with a minimum of three features retained per dimension.

#### 2.5.1 Leave-One-Stream-Out Ablation

To quantify each stream’s contribution to predictive performance, 3 ablation versions (*−S_Geno←LA_*, *−S_Geno←SDOH_*, *−S_LA←SDOH_*) are trained independently. In each version, the pooled output of the disabled stream is zeroed immediately before the softmax gate, while the cross-attention and CLS-pooling computations of that stream continued to execute during the forward pass. The stream is therefore functionally silenced, allowing the gate to redistribute weight onto the two surviving streams. Additionally, single-stream models (retaining only *S_Geno←LA_*, *S_Geno←SDOH_*, or *S_LA←SDOH_*) are trained to assess each stream’s standalone predictive capacity as reported in Supplementary Table S2.

#### 2.5.2 Cross-Modal Pair Extraction

Within each *z*, pairs co-occurring across separate feature modalities are defined as cross-modal pairs and summarized by within-fold co-occurrence (co_count, Jaccard) and across-fold frequency (fold_freq). The outer union of Grad × Activation- and SHAP-derived pairs forms a consensus pool from which pairs passing per-method co-occurrence and Jaccard thresholds are routed into two tiers. The interpretability pipeline (Figure 3) aggregates both measurements into a consensus pool, and divides the pool into two evidence tiers based on cross-fold reproducibility. The thresholds applied are listed in Table 2.

**Table 2:**
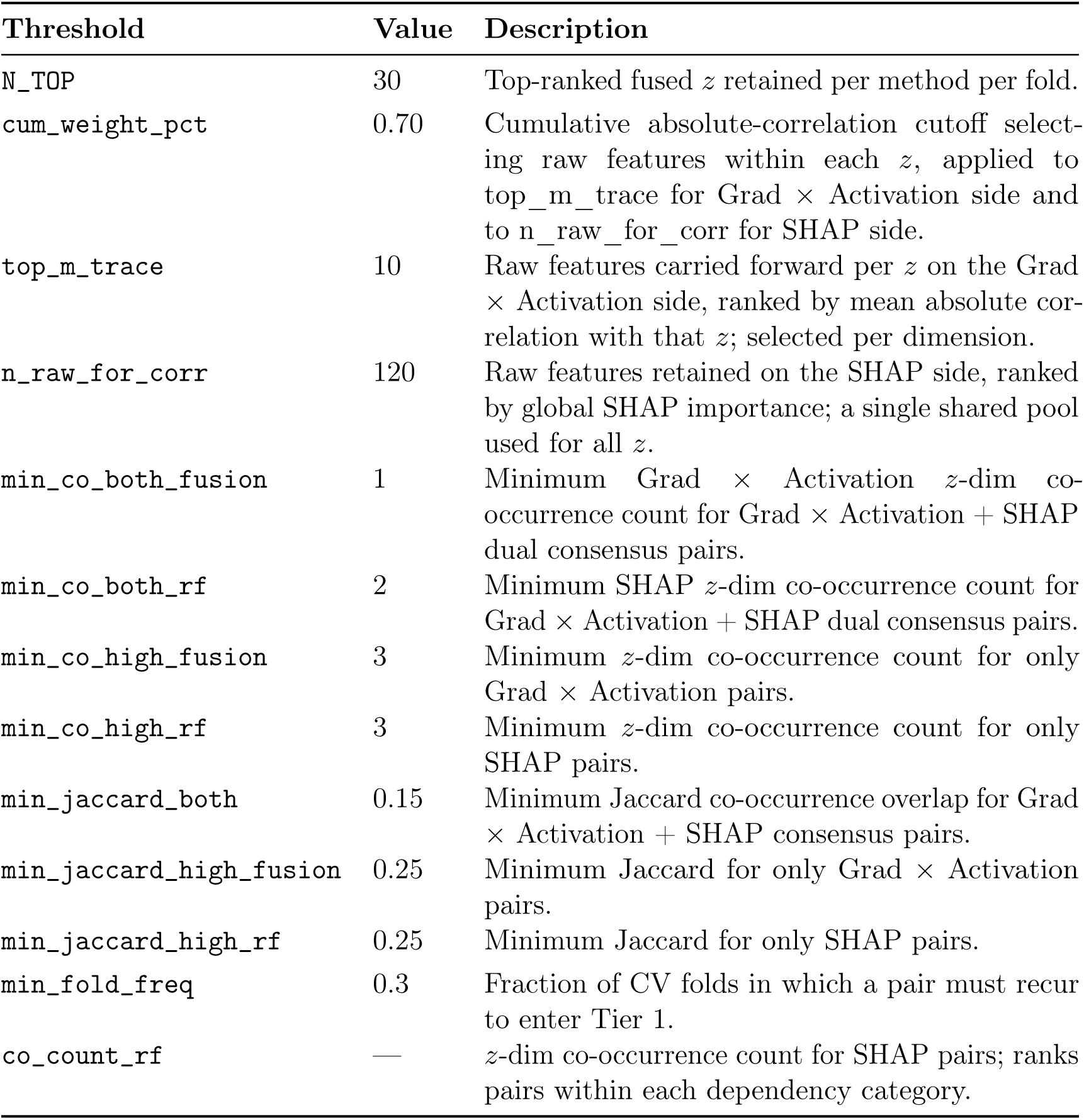
Threshold and ranking parameters of the interpretability pipeline. Discovery parameters (`N_TOP, top_m_trace, n_raw_for_corr, cum_weight_pct`) govern per-fold *z*-dimension and raw-feature retention; the two candidate-set parameters differ between the attribution channels; per-method co-occurrence and Jaccard floors (`min_co_*, min_jaccard_*`) gate the consensus pool; and the `min_fold_freq` gate (≥ 0.3) separates Tier 1 from Tier 2 outputs`. co_count_rf` is a ranking metric, not a filtering threshold.

| Threshold | Value | Description |
| --- | --- | --- |
| `N_TOP` | 30 | Top-ranked fused $z$ retained per method per fold. |
| `cum_weight_pct` | 0.70 | Cumulative absolute-correlation cutoff selecting raw features within each $z$, applied to `top_m_trace` for Grad × Activation side and to `n_raw_for_corr` for SHAP side. |
| `top_m_trace` | 10 | Raw features carried forward per $z$ on the Grad × Activation side, ranked by mean absolute correlation with that $z$; selected per dimension. |
| `n_raw_for_corr` | 120 | Raw features retained on the SHAP side, ranked by global SHAP importance; a single shared pool used for all $z$. |
| `min_co_both_fusion` | 1 | Minimum Grad × Activation $z$-dim co-occurrence count for Grad × Activation + SHAP dual consensus pairs. |
| `min_co_both_rf` | 2 | Minimum SHAP $z$-dim co-occurrence count for Grad × Activation + SHAP dual consensus pairs. |
| `min_co_high_fusion` | 3 | Minimum $z$-dim co-occurrence count for only Grad × Activation pairs. |
| `min_co_high_rf` | 3 | Minimum $z$-dim co-occurrence count for only SHAP pairs. |
| `min_jaccard_both` | 0.15 | Minimum Jaccard co-occurrence overlap for Grad × Activation + SHAP consensus pairs. |
| `min_jaccard_high_fusion` | 0.25 | Minimum Jaccard for only Grad × Activation pairs. |
| `min_jaccard_high_rf` | 0.25 | Minimum Jaccard for only SHAP pairs. |
| `min_fold_freq` | 0.3 | Fraction of CV folds in which a pair must recur to enter Tier 1. |
| `co_count_rf` | — | $z$-dim co-occurrence count for SHAP pairs; ranks pairs within each dependency category. |

Tier 1 retains pairs that meet per-method co-occurrence thresholds and a fold-frequency threshold of 0.3. Tier 2 retains pairs that meet per-method thresholds without the fold-frequency requirement. The two-tier separation reflects empirical heterogeneity in the distribution of cross-modal evidence in this cohort: Geno*←*LA pairs (Tier 1) draw from a large group of features, while cross-modal pairs with four SDOH features (Tier 2) produce within-fold co-occurrence comparable to Tier 1 but rarely clear the fold-frequency gate (Tables 6 and 7). This follows discovery conventions in stability-selection [104].

Within-fold Jaccard and across-fold frequency capture non-redundant aspects of evidence quality (Supplementary Figures S3 and S4. Pairs can have high within-fold jaccard but appear in few folds (strong per-fold signal, fold-distributed routing), or appear consistently across folds with moderate within-fold Jaccard (broadly reproducible, weakly concentrated). The two measurements correlate imperfectly across the consensus pool, supporting the case for treating them as separate axes rather than collapsing them into a single score. The attribution of the fusion architecture contributes to the assignment of levels in a distinctive way. The two attribution channels differ in the size and structure of their candidate pools, and this is reflected in their within-fold Jaccard distributions. On the Grad × Activation side, each latent dimension carries a smaller candidate list, and these lists are largely disjoint across dimensions, so two features rarely co-occur in more than one dimension. On the SHAP side, all latent dimensions draw from a single feature candidate pool, so co-occurrence is common. Supplementary Figure S4 shows this aspect: the median fusion Jaccard is zero across all four configurations, whereas RF Jaccard populates a graded distribution with medians of 0.316-0.333. RF Jaccard is therefore the within-fold metric used to rank pairs above the consensus gates. Additionally, in comparison to Grad × Activation side, higher co-occurrence and Jaccard floors (`min_co_both_rf, min_co_high_rf, min_jaccard_high_rf`) are applied on SHAP side to restrict it to the most recurrent pairs. Ranking within each stream-dependency category in the reported pair tables uses co_count_rf (Table 2).

### 2.6 AoU Independent-Cohort Replication

To assess whether the findings of this study generalize beyond the BIG pediatric case study, the *X-Admix* protocol - modality construction, per-fold feature selection, fusion architecture, training schedule, downstream RF classification, and stream-ablation analyses is repeated end-to-end on an AA adult cohort of 666 participants drawn from the AoU Research Program v7 controlled-tier release (C2022Q4R9) (Supplementary Figure S2).

The AoU analytic cohort comprises 666 self-reported AA adult participants (333 cases, 50%; 333 controls, 50%; 551 female, 82%; median age 59 years, IQR 44–63). Demographic and clinical characteristics of the cohort, alongside the case/control split, are summarized in Table 3. The AoU and BIG cohorts differ in both sex composition and life stage (Tables 1 and 3). Female participants comprised 82% of AoU compared with 49% of BIG, reflecting the underlying composition of AoU enrollment rather than an analytic choice. AoU participants are also substantially older (median age 59 years, IQR 44-63) than BIG participants (median age 16 years, IQR 13-18). Neither sex nor age is included as a covariate in either analysis.

**Table 3:** Demographic and clinical characteristics of the AoU Cohort.

| Characteristics | AoU Cohort |  |  |
| --- | --- | --- | --- |
|  | Overall | Cases | Controls |
| Patients, n (%) | 666 (100) | 333 (50) | 333 (50) |
| Female, n (%) | 551 (82) | 275 (50) | 276 (50) |
| Age, median (IQR) | 59 (44–63) | 59 (44–63) | 59 (44–63) |

The BIG inclusion and ICS non-response criteria (Section 3.1) are applied identically to AoU adult participants: asthma diagnosis by ICD-9 493.x or ICD-10 J45.x, inhaled-corticosteroid prescription, availability of host-genome sequencing, and an emergency-department visit or hospitalization for asthma exacerbation within 24 months of the initial ICS prescription. AoU controls are drawn from participants meeting all inclusion criteria except the exacerbation outcome and are 1:1 age/sex matched to cases (*n* = 333 per arm; Supplementary Figure S2).

AoU participants were sequenced under the AoU short-read whole-genome sequencing (WGS) pipeline, whereas BIG participants were sequenced via WES on Illumina NovaSeq 6000. To make the variant spaces comparable, the AoU WGS variant matrix is restricted to variants overlapping the BIG WES capture region, and the curated neuroimmune and asthma-pathway gene set used to restrict BIG LA candidates (Section 2.2) is applied to the AoU LA features in the same manner. The per-fold selection procedure (Section 2.2) is then applied to this exome-restricted, curated AoU variant set. Per-fold AoU genotype and LA pool sizes are reported in Supplementary Table S1. Genotype and LA selection signal in AoU for the selection procedure is near chance, and per-fold pool sizes were fixed at the smallest set retaining the selection procedure’s peak performance within the training partition of each fold.

The AoU Controlled Tier release provides a 3-digit ZIP-code-level geocoded SDOH panel, spatially coarser than the census-tract-level panel used in BIG. The AoU SDOH candidates are passed through the same per-fold selection procedure used for the other modalities (Section 2.2), which narrowed the panel to five variables retained consistently across all folds: percent vacant housing, median household income, percent with at least a high-school education, percent below the federal poverty line, and percent receiving public-assistance income.

All other architectural and training choices, including pairwise cross-attention streams, gated fusion, MLP head, RF downstream classifier, AdamW with cosine annealing, and 10-fold stratified outer CV, are held identical to the BIG configuration, with one exception. The W&B Bayesian hyperparameter tuning objective differed between cohorts: the BIG sweep optimizes the inner-fold ROC-AUC penalized by its across-split standard deviation (AUC_inner_ *−* 0.5 × SD), whereas the AoU sweep optimizes the inner-fold AUC_inner_ directly, since the AoU cohort is roughly 3x the size of BIG and yielded sufficiently stable inner-fold estimates to render the variance penalty unnecessary.

## 3 Results & Discussion

### 3.1 Predictive performance (BIG)

*S_integrated_* achieves a mean ROC AUC of 0.771 *±* 0.072 across 10-fold CV on the BIG pediatric cohort (Table 4). Among leave-one-stream-out pipelines, *−S_Geno←LA_* produces the highest two-stream AUC (0.779 *±* 0.053), matching *S_integrated_* to within the fold-to-fold SD; *−S_LA←SDOH_* produced a modest decrement (0.743 *±* 0.069); and *−S_Geno←SDOH_* produces the largest decrement (0.656 *±* 0.120). Single-stream pipelines confirm that *S_Geno←SDOH_* alone carried the strongest standalone predictive signal (0.748 *±* 0.173), albeit with substantially higher fold-to-fold SD than any multi-stream configuration (Supplementary Table S2). Gate weights remain approximately uniform across all configurations (*≈* 0.333 per stream), indicating that the model absorbs stream ablation within the attention layers rather than at the gate. These results identify *S_Geno←SDOH_* as the highest-impact stream and *S_Geno←LA_* as the most redundant relative to the *S_integrated_* baseline in terms of predictive performance in this cohort.

**Table 4:** Performance of different model configurations on BIG pediatric cohort. Values are reported as mean *±* SD across folds.

| Model Configuration | ROC AUC | Precision | Recall |
| --- | --- | --- | --- |
| $-S_{\text{Geno} \leftarrow \text{SDOH}}$ | $0.656 \pm 0.120$ | $0.652 \pm 0.113$ | $0.657 \pm 0.122$ |
| $-S_{\text{LA} \leftarrow \text{SDOH}}$ | $0.743 \pm 0.069$ | $0.706 \pm 0.078$ | $0.697 \pm 0.108$ |
| $-S_{\text{Geno} \leftarrow \text{LA}}$ | $0.779 \pm 0.053$ | $0.723 \pm 0.104$ | $0.722 \pm 0.090$ |
| $S_{\text{integrated}}$ | $0.771 \pm 0.072$ | $0.703 \pm 0.080$ | $0.739 \pm 0.136$ |

### 3.2 Classical Interaction Baselines

To benchmark *S_integrated_* against classical interaction modeling under the same 10-fold CV splits, we construct three ridge logistic regression baselines on the BIG cohort. These baselines are as follows: M1 includes only the feature separately per modality (Geno, LA, and SDOH features); M2 retains only the three feature interaction terms (Geno *←* LA, Geno *←* SDOH, LA *←* SDOH) without respective features separately, mirroring the interaction-only architecture that *S_integrated_* pipeline captures; and M3 combines mM1 + M2. All baselines used *L*2-penalized logistic regression (*C* = 1.0) on standardized features (Section 2.2). M1 yield AUC 0.535 *±* 0.114; M2 yields 0.558 *±* 0.134; and M3 yields 0.566 *±* 0.133. The baselines are performed at or near chance on the BIG cohort. This contrast indicates that the improvement is not attributable to multimodal inclusion alone: the tested additive and prespecified product-term models contain the same modalities but do not learn the conditional, directional representations captured by X-Admix.

### 3.3 Pair Discovery

We identify, in Section 2.5.2, cross-modal pairs as features from different modalities that co-occur in shared *z* of the fused representation using a two-method consensus approach.

*S_integrated_* produces 932 consensus cross-modal pairs. Of the 932 *S_integrated_* consensus pairs, 632 (67.8%) are Geno *←* LA, 273 (29.3%) are LA *←* SDOH, and 27 (2.9%) are Geno *←* SDOH. The predominance of Geno *←* LA interactions indicates that the primary cross-modal signal encoded by *S_integrated_* in this cohort is the modulation of genetic-variant effects by LA context.

Beyond predictive ablation mentioned in Section 2.5.1, the same leave-one-stream-out ablations are re-analyzed through the full interpretability pipeline to classify the 932 *S_integrated_* consensus pairs by their dependence on individual cross-attention streams. Note that disabling a stream does not remove any modality from the model inputs. In *−S_Geno←LA_*, for example, genotype and LA features both remain available as inputs and simply cannot attend to each other through the *S_Geno←LA_* stream. Cross-modal *Geno ← LA* pairs can nonetheless still emerge in this setting, because a cross-modal pair is defined by the joint correlation of a feature from each modality with a shared fused latent dimension rather than by an explicit attention stream between the two features. Since genotype and LA are statistically dependent in admixed individuals, a genotype feature and an LA feature may co-embed in the same latent dimension through input-level correlation alone, even when the direct *S_Geno←LA_* stream is removed. This is particularly important because *S_Geno←LA_* stream carries above-chance signal on its own (Supplementary Table S2), but this signal is recoverable without the dedicated *S_Geno←LA_* stream. The pipeline compensates through the surviving streams and through the input-level dependence between genotype and LA.

To characterize how cross-modal pairs depend on each stream, we remove each single stream in turn and record whether each of the 932 *S_integrated_* consensus pairs still pass the consensus thresholds. A pair that no longer pass once a given stream was removed is classified as dependent on that stream (Figure 4a). Stream independent pairs (137; 14.7%), pairs that survived all three ablations, form a methodologically central subset representing cross-modal evidence that the model recovers regardless of which pairwise route is available. Among single-stream-conditional pairs (267; 28.7%), *S_Geno←LA_* dependent pairs are 86 (9.2%), *S_Geno←SDOH_* dependent are 108 (11.6%), and *S_LA←SDOH_* dependent are 73 (7.8%), indicating that no individual stream is uniquely required to recover a dominant fraction of the consensus pool. The multi-stream dependent category (312; 33.5%), pairs lost in two of the three single-stream ablations, is the largest overall, followed by all-stream dependent pairs (216; 23.2%), pairs lost under every single-stream ablation and therefore reproducible only when the full *S_integrated_*pipeline is retained.

**Figure 4:**
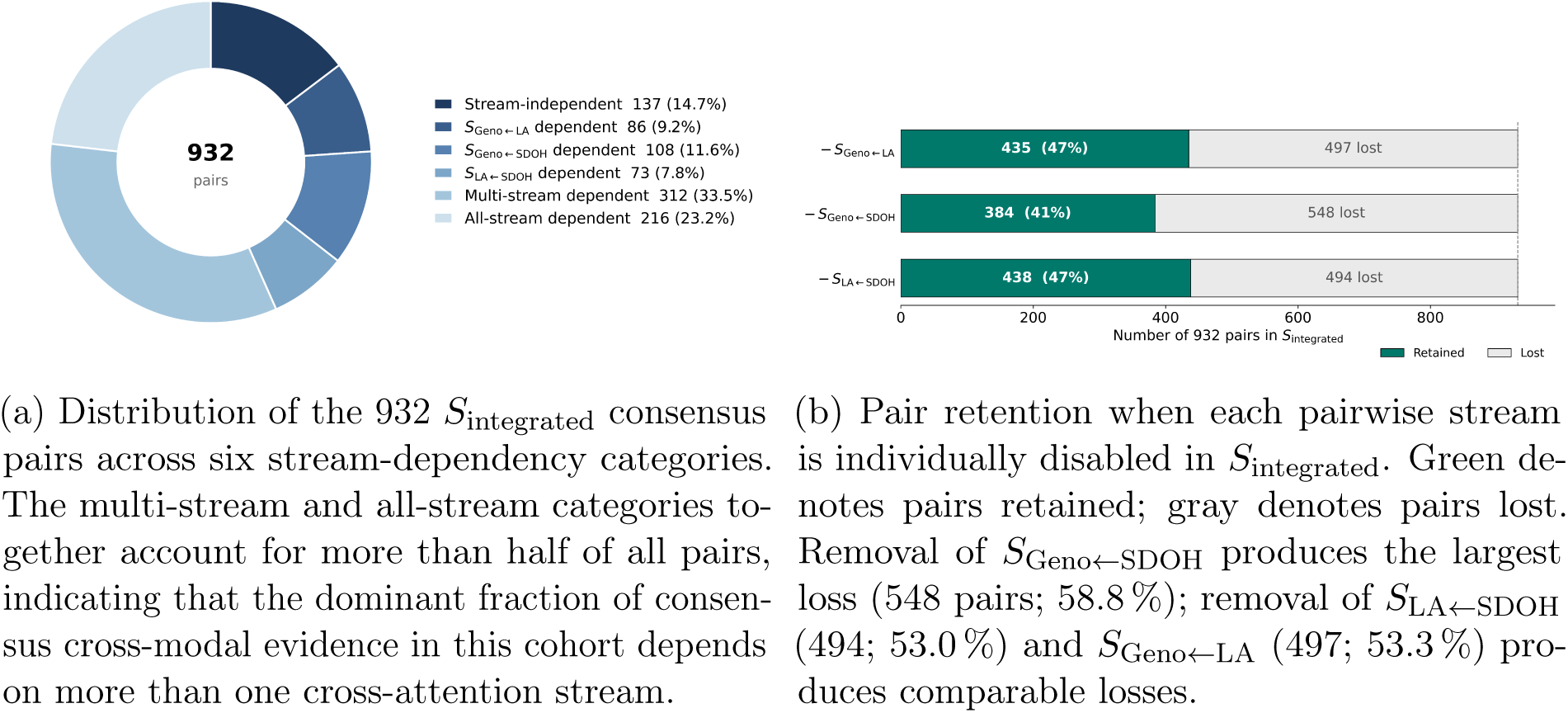
Composition and architectural dependence of consensus cross-modal pairs in *S_integrated_* pipeline.

Representative pairs from each dependency category for Tier 1 and Tier 2 are reported in Tables 6 and 7. Tier 1 is dominated by PAX8 as the genotype-side anchor across the stream independent, *S_Geno←LA_* dependent, and *S_Geno←SDOH_* dependent categories, a recurrence consistent with PAX8 contributing the most cross-fold-reproducible genotype signal in this cohort, with CWF19L2, GALNT10, MYO16, and ABCA4 emerging as additional anchors in the multi-stream and all-stream categories. Tier 2 is dominated by PM2.5 as the SDOH-side anchor, particularly in the stream independent (paired with LOC105375131 and ABCA4) and *S_Geno←SDOH_* dependent categories (paired with PAX8, MYO16, GALNT10, UBR5), reflecting the pre-eminence of ambient air-quality exposure among the four SDOH variables used as inputs. Ancestry-side partners in Tier 2 (LA segments rather than genes) appear concentrated in the multi-stream and all-stream categories, consistent with LA *←* SDOH pairs requiring the integrated *S_integrated_* architecture for stable recovery (Figure 4a; Supplementary Figure S5).

To assess whether ablation altered per-pair quality, we compare the within-fold Jaccard overlap of the two features in each consensus pair across pipelines (Supplementary Figure S4). Median RF Jaccard among retained pairs is essentially preserved under single-stream ablation (0.333 in *S_integrated_*; 0.316 under *−S_Geno←LA_*; 0.333 under both *−S_Geno←SDOH_* and *−S_LA←SDOH_*), with a small reduction limited to *−S_Geno←LA_*. The slight degradation under *−S_Geno←LA_*, taken together with the asymmetric Geno *←* LA pair loss under the same ablation (Supplementary Figure S5), is consistent with the dedicated genotype-ancestry stream contributing modestly to co-embedding concentration without uniquely determining it. The filtered top pairs surviving thresholds change substantially across ablations (Supplementary Figure S5), but the within-pair latent overlap of the pairs that remain is approximately stream-invariant. Because this comparison is conditioned on the pairs each pipeline retains, it shows that ablation does not reduce the co-embedding overlap of surviving pairs; what changes under ablation is which pairs survive.

Geno *←* LA pairs predominate in the consensus pool even though *S_Geno←LA_* contributes little unique predictive signal. The stream’s predictive redundancy is evident in the low AUC cost of removing it (Table 4), while the abundance of Geno *←* LA pairs reflects how consistently genotype and LA features share a fold-stable latent dimension. Predictive and representational importance are therefore not equivalent: a stream can be redundant for aggregate AUC while still shaping which biologically relevant cross-modal relationships the model preserves.

### 3.4 Stream-level Dependency

Stratifying the 932 pairs by stream-dependency category (Section 3.3) reveals a graded architecture of cross-modal evidence rather than a uniform pool. In Tier 1 (Geno *←* LA pairs), the stream-independent category is anchored almost exclusively by *PAX8* paired with a small LA-side panel (*ARHGAP24*, *LTA*, *PSMA6*, *MAPK10*), identifying a *PAX8* -centered Geno *←* LA signal recovered irrespective of route. Single-stream dependent categories partition by the route uniquely require: *PAX8* -*TUBB3*, *PAX8* - *KIF5B*, and *PAX8* -*ITPR1* require *S*_Geno_*_←_*_LA_; *CWF19L2* -*MIR634*, *ABCA4* -*PSMB4*, and *GALNT10* -*IFNGR1* require *S*_LA_*_←_*_SDOH_ despite being Geno *←* LA pairs, indicating that LA features can carry SDOH-routed context into Geno *←* LA co-embedding; and *PAX8* - *PRKCA*, *CWF19L2* -*PSMA6*, and *MYO16* -*PSMA6* depend on *S*_Geno_*_←_*_SDOH_, the same indirect routing in the opposite direction (consistent with the Geno *←* LA turnover pattern in Figure 4b). Multi-stream-dependent pairs (*MYO16* -*PRKCA*, *PAX8* -*CSF1*) and all-stream-dependent pairs (*GALNT10* -*PSMA6*, *CWF19L2* -*ERCC2*, *GALNT10* -*ITPR2*) form the most architecturally coupled subset, requiring the full *S*_integrated_ stream geometry and lost under any single-stream ablation.

Tier 2 (SDOH-based pairs) carries the same graded structure with PM_2.5_ as the dominant SDOH-side anchor. PM_2.5_ paired with *LOC105375131* and *ABCA4* defines a stream-independent core of ambient-air-quality-by-genotype evidence recoverable through any route. The *S*_Geno_*_←_*_SDOH_ dependent block is distinctive: PM_2.5_ and SVI pair with the same geno-side anchors that dominate Tier 1 (*PAX8*, *MYO16*, *GALNT10*, *UBR5*), identifying the direct *S*_Geno_*_←_*_SDOH_ stream as the primary route concentrating ambient-exposure-by-genotype evidence. The all-stream-dependent block is instead dominated by PM_2.5_ paired with LA partners (*ARHGAP24*, *MAPK10*, *PRKCA*, *ITPR2*, *PSMC1*), identifying ancestry-by-exposure interactions as the cross-modal evidence requiring the full *S*_integrated_ architecture and not reproducible under any two-stream configuration. Note that the LA *←* SDOH co-embedding is dependent only under the full *S*_integrated_ framework.

Pathway enrichment across the six stream-dependency categories is computed with Enrichr [94] against seven libraries (KEGG, Reactome, GO Biological Process, MSigDB Hallmark, WikiPathways, BioPlanet, and DisGeNET). Because candidate genes are drawn from a neuro-immune panel, terms recurring across all categories reflect the composition of the candidate set and are not interpreted; the informative comparison is between categories, which share that panel and differ only in routing. *S*_Geno_*_←_*_LA_-dependent pairs enrich for Toll-like receptor 3 and endolysosomal TLR signaling and for dendritic-cell cytokine production, implicating ancestry-modulated innate sensing; *S*_Geno_*_←_*_SDOH_-dependent pairs enrich for Th1/Th2 differentiation, Fc*ε*RI signaling, ErbB and JNK/TAK1 cascades, and AGE-RAGE signaling, consistent with adaptive allergic-immune programs routed through the *S*_Geno_*_←_*_SDOH_ stream; and all-stream-dependent pairs enrich for core transcription and DNA-repair processes (RNA polymerase II initiation/elongation, GG-NER, iron-sulfur cluster assembly), biology retained only under *S*_integrated_ and lost under every single-stream ablation. Most directly relevant to the clinical phenotype, Tier 2 all-stream-dependent pairs surface *Occupational Asthma* (DisGeNET), inflammatory-mediator regulation of TRP channels, and Fc*ε*RI-mediated MAPK activation. These category-specific enrichments define an environmental-exposure-by-allergic-immune signature that separates from the other categories only when genotype, LA, and SDOH are jointly modeled, providing candidate mechanistic context for the cross-modal predictive signal *S*_integrated_ captures. Thus, the framework provides more than separate feature-importance lists: it identifies which genetic, LA, and exposure features are connected, the information route supporting each relationship, and whether that relationship depends on retaining a particular cross-attention stream.

### 3.5 AoU Performance

To assess whether the predictive-load dissociation observed in BIG generalizes to a larger admixed cohort, we train and evaluate *S_integrated_* and the three leave-one-stream-out ablations on the AoU adult cohort under the same 10-fold CV protocol (Table 8).

As a single-modality pipeline on the AoU cohort, an SDOH-only pipeline, following the same feature selection pipelines mentioned in Section 2.2, achieves ROC AUC 0.727 *±* 0.071, balanced precision 0.694 *±* 0.069, and balanced recall 0.689 *±* 0.064, comparable within fold-to-fold SD of the best AoU configuration (*S*_integrated_, 0.736 *±* 0.052). By contrast, single-modality genotype-only and LA-only configurations on the same AoU folds yielded metrics at or near chance (*≈* 0.50), confirming that area-level SDOH is the major modality carrying predictive signal in this adult cohort. This contrast is consistent with a known biological divergence: pediatric asthma is more genetically driven, whereas adult asthma is phenotypically heterogeneous and shaped by cumulative environmental and socioeconomic exposures, likely contributing to the stronger genotype-stream signal observed in BIG relative to stronger SDOH-stream signal AoU [105, 106].

Applying the interpretability pipeline to AoU yields substantially fewer top cross-modal pairs, with only two all-stream-dependent gene pairs surviving as Tier 1 and no pairs surviving as Tier 2. The consensus gates retain a pair only when its two features co-occur in at least three shared latent dimensions and recur across folds, so the size of the recovered pool reflects how concentrated and how reproducible the cross-modal co-embeddings are. Both properties are weaker in AoU, and they trace to the same cause. Genotype-only and LA-only pipelines perform near chance in this cohort, consistent with adult asthma being shaped by cumulative environmental and socioeconomic exposure rather than the more genetically driven pediatric phenotype [105, 106]. With little stable genotype or LA signal to lock onto, the per-fold feature selection becomes unstable, returning genotype and LA pools that are far smaller and more variable than in BIG (Supplementary Table S1). The smaller pools give features fewer chances to reach the co-occurrence threshold, while the fold-to-fold variability keeps pairs from recurring often enough to meet the fold-frequency gate. The same instability explains the empty Tier 2 because cross-modal pairing requires a stable genotype or ancestry partner, even though SDOH predicts well on its own. Therefore, the replication between cohorts rests on the stream-level predictive pattern rather than on cross-modal pair identification.

The broader pattern is consistent across the two cohorts: the genotype-ancestry stream is architecturally replaceable on AUC, while the two SDOH-based streams carry the load-bearing predictive signal. *S_integrated_* achieves AUC 0.736 *±* 0.052; *−S_Geno←LA_* achieves 0.735 *±* 0.036; and the two SDOH-based ablations produce larger decrements (*−S_Geno←SDOH_* = 0.710 *±* 0.047; *−S_LA←SDOH_* = 0.711 *±* 0.057). AoU also replicates BIG on the point that removing *S_Geno←LA_* does not measurably change predictive performance in either cohort. The BIG-specific performance in general specially the difference between the performance metrics of the two SDOH-based ablations (Section 3.1) does not replicate at the same magnitude in AoU.

### 3.6 Limitations and Future Work

Both cohorts remain modest for attention-based representation learning, especially BIG (n=240); nested cross-validation and the two-stage encoder–RF design reduce but do not eliminate overfitting risk. Larger ancestry-aligned cohorts are needed to assess robustness and transportability. Moreover, fold-wise standard deviation measures variability across data splits rather than patient-level predictive uncertainty. Model calibration and individual-level uncertainty were not evaluated and should be assessed before clinical use. Similarly, the inability to track medication adherence may have conflated non-compliance with treatment failure, and incorporating adherence monitoring in future studies would make target variable more robust.

Additionally, the BIG-specific asymmetry between SDOH-based ablations (*−S*_Geno_*_←_*_SDOH_ = 0.656 vs. *−S*_LA_*_←_*_SDOH_ = 0.743) collapses in AoU (0.710 vs. 0.711), plausibly reflecting differences in life stage (pediatric vs. adult median age), geographic exposure homogeneity (Memphis vs. nationally distributed), and statistical resolution (*n* = 240 vs. 666); disentangling these requires larger, developmentally heterogeneous admixed cohorts.

The SDOH panels also differ structurally: BIG employs four Census tract level geocoded variables (PM_2.5_, SVI, ADI, urban/rural status) whereas AoU relies on five 3-digit ZIP coded socioeconomic indices with no air-quality measure, so PM_2.5_-based Tier 2 pairs dominating BIG (Table 7) cannot be replicated in AoU by construction. Local ancestry is also encoded as a binary AFR-presence indicator per variant, which collapses one and two AFR haplotype copies into a single category thus discards dosage information; a 0/1/2 copy-number encoding should be evaluated in future work.

Three aspects of the interpretability pipeline warrant explicit acknowledgment. First, the latent-to-feature tracing procedure draws both attribution layers from the same trained model, introducing a degree of circularity; we therefore treat the resulting pair sets as model-prioritized candidates for downstream validation rather than as independently confirmed interactions. Second, the cross-modal pair extraction procedure is a prioritization heuristic based on attribution concordance, not a formal statistical interaction test; reported pairs identify features that the model co-embeds in top important latent dimensions and should be read as candidates for replication validation rather than as confirmed biological interactions.

The two attribution channels are not independent. Grad × Activation is computed through the MLP head and SHAP through the downstream RF, but both operate on the latent space *z* of the same trained encoder, and both link a dimension to raw features by the same correlation. They differ only in how the candidate set of raw features for that correlation is formed (Section 2.5). Thus, the 932 *S*_integrated_ consensus pairs and their pathway enrichment should be treated as model-prioritized candidates rather than validated interactions.

A key direction for future work is to test whether the recovered Geno *←* LA structure reflects locus-specific cross-modal signal or shared global ancestry. This can be approached by accounting for global ancestry in the framework. Another direction is to subject the surfaced candidate pairs to functional or external validation, the step required to move them from model-prioritized hypotheses toward confirmed cross-modal interactions.

### 3.7 Biological and clinical significance

Taken together, these results point to a specific biological reading of why patients with similar diagnoses, or even similar genotypes, differ in their response to inhaled corticosteroids: the effect of a variant need not be fixed. The same allele may carry different phenotypic information depending on the ancestral origin of the genomic segment on which it sits and on the environmental and SDOH context of the individual who carries it. The strongest category-specific signal we recover is consistent with this reading—Th1/Th2 differentiation, a central axis of allergic-immune programming, emerges specifically through the genotype-conditioned-on-SDOH (Geno*←*SDOH) route, as expected if environmental exposure modifies genotype-linked immune pathways rather than acting on them independently. Ambient air-quality exposure (PM2.5) similarly links exposure context to genotype and LA representations. The contribution to asthma is therefore two-fold: improved prediction of a heterogeneous treatment-response phenotype in an underrepresented population, and the organization of genotype, LA, and SDOH signal into route-annotated, testable hypotheses about that heterogeneity. We report these relationships as model-prioritized candidates for statistical and functional validation, not as established interactions.

## 4 Conclusion

To the best of our knowledge, X-Admix is the first framework to jointly represent genotype, LA, and SDOH through cross-attention in admixed cohorts, and it yields two principal findings. First, the integrated three-stream model predicts ICS non-response with a mean AUC of 0.771 *±* 0.072 under 10-fold cross-validation in a pediatric AA severe-asthma cohort, well above classical ridge-logistic baselines that perform near chance (AUC *≤* 0.566; Tables 4 and 5). This pattern replicates in an independent 666-participant AoU adult cohort, where *S*_integrated_ and *−S*_Geno_*_←_*_LA_ yield nearly identical AUC (Tables 4 and 8). Second, a stream’s contribution to interpretation is separable from its contribution to prediction: removing *S*_Geno_*_←_*_LA_ leaves AUC unchanged yet changes the top cross-modal pairs recovered (Figure 4a), so a stream redundant for prediction can still define the candidate interactions carried forward for downstream analysis. The contribution is therefore not merely multimodal inclusion, but the learning of directional, conditional representations across genotype, LA, and SDOH. More broadly, X-Admix organizes genetic, LA, and social-environmental information into testable hypotheses about why patients with similar diagnoses or genotypes may respond differently to ICS. Together, these results yield an interpretable catalog of candidate cross-modal interactions underlying ICS non-response, which we report as model-prioritized hypotheses for validation in independent ancestry-aligned cohorts.

**Table 5:** Performance of classical ridge logistic regression baselines on BIG pediatric cohort. Values are reported as mean *±* SD across 10-fold CV.

| Model Configuration | ROC AUC |
| --- | --- |
| M1: main effects only | 0.535 ± 0.114 |
| M2: all interactions only | 0.558 ± 0.134 |
| M3: M1 + M2 | 0.566 ± 0.133 |

**Table 6:**
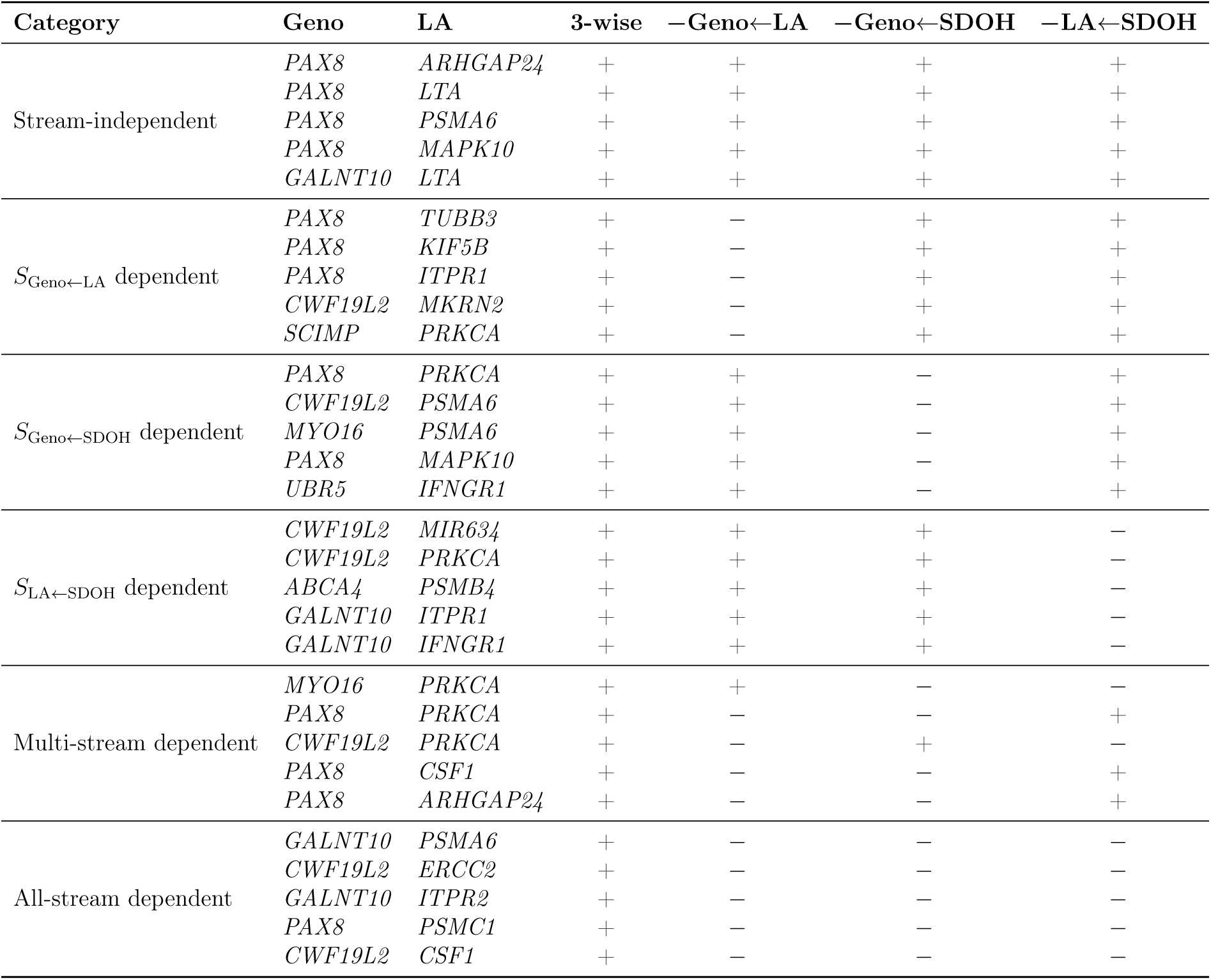
Top-ranked Tier 1 (Geno–LA) cross-attention attribution pairs per dependency category. All pairs consist of an Geno (feature 1) and an LA (feature 2) features; modality columns are omitted for brevity. The *Category*column indicates which attention stream(s) are required for the pair to appear: *Stream independent* pairs persist across all ablations; *All-stream dependent* pairs vanish when any single stream is removed. Columns *S*_Geno_*_←_*_LA_, *S*_Geno_*_←_*_SDOH_, and *S*_LA_*_←_*_SDOH_ report pair presence (+) or absence (*−*) in each single-stream ablation.

**Table 7:** Top-ranked Tier 2 (SDOH–Geno/LA) cross-attention attribution pairs per dependency category. All pairs have an SDOH feature as feature 1; the modality column for feature 1 is omitted for brevity. feature 1 values (PM2.5, Urban, SVI, ADI) are SDOH features; feature 2 values are gene symbols for Geno or LA features.

| Category | SDOH | Modality 2 | Feature 2 | 3-wise | -Geno←LA | -Geno←SDOH | -LA←SDOH |
| --- | --- | --- | --- | --- | --- | --- | --- |
| Stream-independent | PM2.5 | Geno | *LOC105375131* | + | + | + | + |
|  | PM2.5 | Geno | *ABCA4* | + | + | + | + |
|  | Urban | Geno | *LOC105375131* | + | + | + | + |
|  | SVI | Geno | *LOC105375131* | + | + | + | + |
| $S_{\text{Geno} \leftarrow \text{LA}}$ dependent | PM2.5 | Geno | *SCIMP* | + | - | + | + |
|  | SVI | LA | *PSMB4* | + | - | + | + |
| $S_{\text{Geno} \leftarrow \text{SDOH}}$ dependent | PM2.5 | Geno | *PAX8* | + | + | - | + |
|  | PM2.5 | Geno | *MYO16* | + | + | - | + |
|  | PM2.5 | Geno | *GALNT10* | + | + | - | + |
|  | PM2.5 | Geno | *UBR5* | + | + | - | + |
|  | SVI | Geno | *MYO16* | + | + | - | + |
| $S_{\text{LA} \leftarrow \text{SDOH}}$ dependent | SVI | Geno | *ABCA4* | + | + | + | - |
| Multi-stream dependent | ADI | Geno | *PAX8* | + | - | - | + |
|  | PM2.5 | Geno | *CWF19L2* | + | + | - | - |
|  | Urban | LA | *ARHGAP24* | + | + | - | - |
|  | ADI | LA | *LTA* | + | + | - | - |
|  | Urban | Geno | *GALNT10* | + | - | - | + |
| All-stream dependent | PM2.5 | LA | *ARHGAP24* | + | - | - | - |
|  | PM2.5 | LA | *MAPK10* | + | - | - | - |
|  | PM2.5 | LA | *PRKCA* | + | - | - | - |
|  | PM2.5 | LA | *ITPR2* | + | - | - | - |
|  | PM2.5 | LA | *PSMC1* | + | - | - | - |

**Table 8:** Performance of different model configurations on AoU adult cohort. Values are reported as mean *±* SD across folds. Best value in each metric is shown in bold.

| Model Configuration | ROC AUC | Precision | Recall |
| --- | --- | --- | --- |
| $-S_{\text{Geno} \leftarrow \text{SDOH}}$ | $0.710 \pm 0.047$ | $0.658 \pm 0.049$ | $0.691 \pm 0.107$ |
| $-S_{\text{LA} \leftarrow \text{SDOH}}$ | $0.711 \pm 0.057$ | $0.663 \pm 0.071$ | $0.672 \pm 0.103$ |
| $-S_{\text{Geno} \leftarrow \text{LA}}$ | $0.735 \pm 0.036$ | $0.679 \pm 0.042$ | $0.733 \pm 0.090$ |
| $S_{\text{integrated}}$ | $0.736 \pm 0.052$ | $0.680 \pm 0.054$ | $0.705 \pm 0.088$ |

## Data Availability

The BIG Initiative data analyzed here were collected under IRB #22-09164-NHSR and contain protected genomic and clinical information. Whole exome sequencing was carried out for all subjects in our biorepository project that has been funded through internal (non-federal) funding sources within our Institution. As a result, we have not deposited the data in dbGaP. However, we have a well developed process that enable investigators to request data. Our consent form indicates, and our Institution requires, that all use of our biorepository data by outside investigators be carried out in the context of a fully collaborative research project. To request data access, please visit: https://uthsc.edu/cbmi/big/. This study used data from the All of Us Research Program’s Controlled Tier Dataset v7 (C2022Q4R9), available to authorized users on the Researcher Workbench.

## Acknowledgements

We thank the Regeneron Genetics Center for their sequencing services. We gratefully acknowledge All of Us participants for their contributions, without whom this research would not have been possible. We also thank the National Institutes of Health’s All of Us Research Program for making available the participant data examined in this study.

## Ethics Statement

The BIG Initiative study was approved by the Institutional Review Board of the University of Tennessee Health Science Center (IRB #22-09164-NHSR). Informed consent was obtained from all participants or their legal guardians in accordance with the Declaration of Helsinki. Analyses of All of Us Research Program data were conducted on the Researcher Workbench under the Program’s Data Use and Registration Agreement using the Controlled Tier Dataset v7 (C2022Q4R9).

## Use of Large Language Models

Large language model-based tools were used for language editing during preparation of this manuscript. All scientific content, analyses, and interpretations were generated and verified by the authors.

## References

[1] J. N. Acosta, G. J. Falcone, P. Rajpurkar, and E. J. Topol. Multimodal biomedical ai. Nat Med, 28(9):1773–1784, 2022. ISSN 1078-8956. doi: 10.1038/s41591-022-01981-2.

[2] Adrienne Kline, Hanyin Wang, Yikuan Li, Saya Dennis, Meghan Hutch, Zhenxing Xu, Fei Wang, Feixiong Cheng, and Yuan Luo. Multimodal machine learning in precision health: A scoping review. NPJ digital medicine, 5(1):171, 2022. ISSN 2398-6352.

[3] S. Steyaert, Y. L. Qiu, Y. Zheng, P. Mukherjee, H. Vogel, and O. Gevaert. Multimodal deep learning to predict prognosis in adult and pediatric brain tumors. Commun Med (Lond), 3(1):44, 2023. ISSN 2730-664x. doi: 10.1038/s43856-023-00276-y.

[4] Jana Lipkova, Richard J Chen, Bowen Chen, Ming Y Lu, Matteo Barbieri, Daniel Shao, Anurag J Vaidya, Chengkuan Chen, Luoting Zhuang, and Drew FK Williamson. Artificial intelligence for multimodal data integration in oncology. Cancer cell, 40 (10):1095–1110, 2022. ISSN 1535-6108.

[5] E. G. Atkinson, A. X. Maihofer, M. Kanai, A. R. Martin, K. J. Karczewski, M. L. Santoro, J. C. Ulirsch, Y. Kamatani, Y. Okada, H. K. Finucane, K. C. Koenen, C. M. Nievergelt, M. J. Daly, and B. M. Neale. Tractor uses local ancestry to enable the inclusion of admixed individuals in gwas and to boost power. Nat Genet, 53(2): 195–204, 2021. ISSN 1061-4036 (Print) 1061-4036. doi: 10.1038/s41588-020-00766-y.

[6] Bogdan Pasaniuc, Noah Zaitlen, Guillaume Lettre, Gary K Chen, Arti Tandon, WH Linda Kao, Ingo Ruczinski, Myriam Fornage, David S Siscovick, and Xiaofeng Zhu. Enhanced statistical tests for gwas in admixed populations: assessment using african americans from care and a breast cancer consortium. PLoS genetics, 7(4):e1001371, 2011. ISSN 1553-7390. URL https://journals.plos.org/plosgenetics/article/file?id=10.1371/journal.pgen.1001371&type=printable.

[7] M. F. Seldin, B. Pasaniuc, and A. L. Price. New approaches to disease mapping in admixed populations. Nat Rev Genet, 12(8):523–8, 2011. ISSN 1471-0056 (Print) 1471-0056. doi: 10.1038/nrg3002.

[8] Brian K Maples, Simon Gravel, Eimear E Kenny, and Carlos D Bustamante. Rfmix: a discriminative modeling approach for rapid and robust local-ancestry inference. The American Journal of Human Genetics, 93(2):278–288, 2013. ISSN 0002-9297. URL https://pmc.ncbi.nlm.nih.gov/articles/PMC3738819/pdf/main.pdf.

[9] Kangcheng Hou, Arjun Bhattacharya, Rachel Mester, Kathryn S Burch, and Bogdan Pasaniuc. On powerful gwas in admixed populations. Nature genetics, 53(12): 1631–1633, 2021. ISSN 1061-4036.

[10] Quan Sun, Andrea RVR Horimoto, Brian Chen, Frank Ockerman, Karen L Mohlke, Elizabeth Blue, Laura M Raffield, and Yun Li. Opportunities and challenges of local ancestry in genetic association analyses. The American Journal of Human Genetics, 112(4):727–740, 2025. ISSN 0002-9297.

[11] Costa Georgantas, Zoltán Kutalik, and Jonas Richiardi. Delphi: Deep learning for polygenic risk prediction. medRxiv, page 2024.04.19.24306079, 2026. doi: 10.1101/2024.04.19.24306079. URL https://www.medrxiv.org/content/medrxiv/early/2026/02/06/2024.04.19.24306079.full.pdf.

[12] A. Singh, C. Alarcon, E. A. Nutescu, T. J. O’Brien, M. Tuck, L. Gong, T. E. Klein, D. O. Meltzer, J. A. Johnson, L. H. Cavallari, and M. A. Perera. Local ancestry informed gwas of warfarin dose requirement in african americans identifies a novel cyp2c19 splice qtl. medRxiv, 2025. doi: 10.1101/2025.03.03.25323247.

[13] Katarzyna Bryc, Eric Y Durand, J Michael Macpherson, David Reich, and Joanna L Mountain. The genetic ancestry of african americans, latinos, and european americans across the united states. The American Journal of Human Genetics, 96(1):37–53, 2015. ISSN 0002-9297.

[14] Tesfaye B Mersha, Ke Qin, Andrew F Beck, Lili Ding, Bin Huang, and Robert S Kahn. Genetic ancestry differences in pediatric asthma readmission are mediated by socioenvironmental factors. Journal of Allergy and Clinical Immunology, 148(5): 1210–1218. e4, 2021. ISSN 0091-6749.

[15] Hari S Iyer, Iona Cheng, Chidinma Opara, Katherine Lin, Nur Zeinomar, Loïc Le Marchand, Lynne Wilkens, Salma Shariff-Marco, David V Conti, and Christopher A Haiman. African genetic ancestry, structural and social determinants of health, and mortality in black adults. JAMA Network Open, 8(5):e2510016, 2025. ISSN 2574-3805.

[16] Heather J Cordell. Detecting gene–gene interactions that underlie human diseases. Nature Reviews Genetics, 10(6):392–404, 2009. ISSN 1471-0056.

[17] Kimberly McAllister, Leah E Mechanic, Christopher Amos, Hugues Aschard, Ian A Blair, Nilanjan Chatterjee, David Conti, W James Gauderman, Li Hsu, and Carolyn M Hutter. Current challenges and new opportunities for gene-environment interaction studies of complex diseases. American journal of epidemiology, 186(7): 753–761, 2017. ISSN 0002-9262.

[18] Duncan Thomas. Gene–environment-wide association studies: emerging approaches. Nature reviews genetics, 11(4):259–272, 2010. ISSN 1471-0056. URL https://www.nature.com/articles/nrg2764.

[19] Alkes L Price, Nick J Patterson, Robert M Plenge, Michael E Weinblatt, Nancy A Shadick, and David Reich. Principal components analysis corrects for stratification in genome-wide association studies. Nature genetics, 38(8):904–909, 2006. ISSN 1061-4036. URL https://www.nature.com/articles/ng1847.

[20] Jian Yang, S Hong Lee, Michael E Goddard, and Peter M Visscher. Gcta: a tool for genome-wide complex trait analysis. The American journal of human genetics, 88 (1):76–82, 2011. ISSN 0002-9297.

[21] Xiang Zhou and Matthew Stephens. Genome-wide efficient mixed-model analysis for association studies. Nature genetics, 44(7):821–824, 2012. ISSN 1061-4036. URL https://www.nature.com/articles/ng.2310.

[22] Po-Ru Loh, Gleb Kichaev, Steven Gazal, Armin P Schoech, and Alkes L Price. Mixed-model association for biobank-scale datasets. Nature genetics, 50(7):906–908, 2018. ISSN 1061-4036. URL https://www.nature.com/articles/s41588-018-0144-6.

[23] Gad Abraham, Adam Kowalczyk, Justin Zobel, and Michael Inouye. Performance and robustness of penalized and unpenalized methods for genetic prediction of complex human disease. Genetic epidemiology, 37(2):184–195, 2013. ISSN 0741-0395.

[24] Florian Privé, Hugues Aschard, and Michael GB Blum. Efficient implementation of penalized regression for genetic risk prediction. Genetics, 212(1):65–74, 2019. ISSN 1943-2631.

[25] Ashish Vaswani, Noam Shazeer, Niki Parmar, Jakob Uszkoreit, Llion Jones, Aidan N Gomez, Łukasz Kaiser, and Illia Polosukhin. Attention is all you need. Advances in neural information processing systems, 30, 2017.

[26] Arsha Nagrani, Shan Yang, Anurag Arnab, Aren Jansen, Cordelia Schmid, and Chen Sun. Attention bottlenecks for multimodal fusion. Advances in neural information processing systems, 34:14200–14213, 2021.

[27] Xuguang Zhou, Chen Chen, Xiaoyi Lv, Enguang Zuo, Min Li, Lijun Wu, Xiaomei Chen, Xue Wu, and Cheng Chen. Cmacf: Transformer-based cross-modal attention cross-fusion model for systemic lupus erythematosus diagnosis combining raman spectroscopy, ftir spectroscopy, and metabolomics. Information Processing & Management, 61(6):103804, 2024. ISSN 0306-4573. doi: 10.1016/j.ipm.2024.103804. URL https://www.sciencedirect.com/science/article/pii/S0306457324001638.

[28] Konstantin Hemker, Nikola Simidjievski, and Mateja Jamnik. Healnet: Multimodal fusion for heterogeneous biomedical data. Advances in Neural Information Processing Systems, 37:64479–64498, 2024.

[29] Sören Richard Stahlschmidt, Benjamin Ulfenborg, and Jane Synnergren. Multimodal deep learning for biomedical data fusion: a review. Briefings in bioinformatics, 23 (2):bbab569, 2022. ISSN 1467-5463.

[30] Richard J Chen, Ming Y Lu, Wei-Hung Weng, Tiffany Y Chen, Drew FK Williamson, Trevor Manz, Maha Shady, and Faisal Mahmood. Multimodal co-attention transformer for survival prediction in gigapixel whole slide images. In Proceedings of the IEEE/CVF international conference on computer vision, pages 4015–4025.

[31] Zinzi D Bailey, Nancy Krieger, Madina Agénor, Jasmine Graves, Natalia Linos, and Mary T Bassett. Structural racism and health inequities in the usa: evidence and interventions. The lancet, 389(10077):1453–1463, 2017. ISSN 0140-6736.

[32] Adali Martinez, Rosemarie de la Rosa, Mahasin Mujahid, and Neeta Thakur. Structural racism and its pathways to asthma and atopic dermatitis. Journal of Allergy and Clinical Immunology, 148(5):1112–1120, 2021. ISSN 0091-6749.

[33] David R Williams, Jourdyn A Lawrence, and Brigette A Davis. Racism and health: evidence and needed research. Annual review of public health, 40(1):105–125, 2019. ISSN 0163-7525.

[34] Luisa N Borrell, Jennifer R Elhawary, Elena Fuentes-Afflick, Jonathan Witonsky, Nirav Bhakta, Alan HB Wu, Kirsten Bibbins-Domingo, José R Rodríguez-Santana, Michael A Lenoir, and James R Gavin III. Race and genetic ancestry in medicine—a time for reckoning with racism, 2021.

[35] Anna CF Lewis, Santiago J Molina, Paul S Appelbaum, Bege Dauda, Anna Di Rienzo, Agustin Fuentes, Stephanie M Fullerton, Nanibaa’A Garrison, Nayanika Ghosh, and Evelynn M Hammonds. Getting genetic ancestry right for science and society. Science, 376(6590):250–252, 2022. ISSN 0036-8075.

[36] Jessica P Cerdeña, Vanessa Grubbs, and Amy L Non. Genomic supremacy: the harm of conflating genetic ancestry and race. Human Genomics, 16(1):18, 2022. ISSN 1479-7364. URL https://link.springer.com/content/pdf/10.1186/s40246-022-00391-2.pdf.

[37] Joshua M Galanter, Christopher R Gignoux, Sam S Oh, Dara Torgerson, Maria Pino-Yanes, Neeta Thakur, Celeste Eng, Donglei Hu, Scott Huntsman, and Harold J Farber. Differential methylation between ethnic sub-groups reflects the effect of genetic ancestry and environmental exposures. elife, 6:e20532, 2017. ISSN 2050-084X.

[38] Matteo D’Antonio, Wilfredo G. Gonzalez Rivera, Robert A Greenes, Melissa Gymrek, and Kelly A Frazer. A highly accurate risk factor-based xgboost multiethnic model for identifying patients with skin cancer. Nature Communications, 16(1):9542, 2025. ISSN 2041-1723.

[39] Natalia Hernandez-Pacheco, Niloufar Farzan, Ben Francis, Leila Karimi, Katja Repnik, Susanne J Vijverberg, Patricia Soares, Maximilian Schieck, Mario Gorenjak, and Erick Forno. Genome-wide association study of inhaled corticosteroid response in admixed children with asthma. Clinical & Experimental Allergy, 49(6):789–798, 2019. ISSN 0954-7894.

[40] Erick Forno and Juan C Celedón. Health disparities in asthma, 2012.

[41] Karen E Wells, Sonia Cajigal, Edward L Peterson, Brian K Ahmedani, Rajesh Kumar, David E Lanfear, Esteban G Burchard, and L Keoki Williams. Assessing differences in inhaled corticosteroid response by self-reported race-ethnicity and genetic ancestry among asthmatic subjects. Journal of allergy and clinical immunology, 137(5): 1364–1369. e2, 2016. ISSN 0091-6749.

[42] Monica H Wojcik, Talia S Schwartz, Katri E Thiele, Heather Paterson, Rachel Stadelmaier, Thomas E Mullen, Grace E VanNoy, Casie A Genetti, Jill A Madden, and Cynthia S Gubbels. Infant mortality: the contribution of genetic disorders. Journal of Perinatology, 39(12):1611–1619, 2019. ISSN 0743-8346. URL https://www.nature.com/articles/s41372-019-0451-5.

[43] Leah E. Mechanic and Carolyn M. Hutter. Gene-Environment Interactions in Human Health, pages 229–249. Springer London, London, 2015. ISBN 978-1-4471-6678-8. doi: 10.1007/978-1-4471-6678-8_10. URL 10.1007/978-1-4471-6678-8_10https://link.springer.com/chapter/10.1007/978-1-4471-6678-8_10.

[44] All of Us Research Program Investigators. The “all of us” research program. New England Journal of Medicine, 381(7):668–676, 2019. ISSN 0028-4793.

[45] AoURPG Investigators. Genomic data in the all of us research program. Nature, 627(8003):340–346, 2024.

[46] Samantha Tesfaye, Robert M Cronin, Maria Lopez-Class, Qingxia Chen, Christopher S Foster, Callie A Gu, Andrew Guide, Robert A Hiatt, Angelica S Johnson, and Christine LM Joseph. Measuring social determinants of health in the all of us research program. Scientific Reports, 14(1):8815, 2024. ISSN 2045-2322. URL https://www.nature.com/articles/s41598-024-57410-6.pdf.

[47] Jing Lei, Max G’Sell, Alessandro Rinaldo, Ryan J. Tibshirani, and Larry Wasserman. Distribution-free predictive inference for regression. Journal of the American Statistical Association, 113(523):1094–1111, 2018. ISSN 0162-1459. doi: 10.1080/01621459.2017.1307116. URL 10.1080/01621459.2017.1307116.

[48] Leo Breiman. Random forests. Machine Learning, 45(1):5–32, 2001. ISSN 1573-0565. doi: 10.1023/A:1010933404324. URL 10.1023/A:1010933404324https://link.springer.com/content/pdf/10.1023/A:1010933404324.pdf.

[49] Galit Shmueli. To explain or to predict? Statistical Science, 25(3):289–310, 2010. ISSN 08834237. URL http://www.jstor.org/stable/41058949.

[50] Laura Toloşi and Thomas Lengauer. Classification with correlated features: unreliability of feature ranking and solutions. Bioinformatics, 27(14):1986–1994, 2011. ISSN 1367-4803. doi: 10.1093/bioinformatics/btr300. URL 10.1093/bioinformatics/btr300.

[51] Hanbin Lee, Moo Hyuk Lee, Kangcheng Hou, Bogdan Pasaniuc, and Buhm Han. Admixed and single-continental genome segments of the same ancestry have distinct linkage disequilibrium patterns. Genome Biology, 26(1):201, 2025. ISSN 1474-760X. doi: 10.1186/s13059-025-03672-w. URL 10.1186/s13059-025-03672-whttps://link.springer.com/content/pdf/10.1186/s13059-025-03672-w.pdf.

[52] Nahian Tahmin, Lokesh K. Chinthala, Franco Leonel Marsico, Silvia Buonaiuto, Akram Mohammed, Annette Carlisle, Yadu Gautam, Vincenza Colonna, Tesfaye B. Mersha, Robert L. Davis, and Anahita Khojandi. Machine learning models incorporating genotype and ancestry improve severe asthma risk prediction. Scientific Reports, 15(1):40243, 2025. ISSN 2045-2322. doi: 10.1038/s41598-025-24080-x. URL 10.1038/s41598-025-24080-xhttps://www.nature.com/articles/s41598-025-24080-x.pdf.

[53] Jonathan Pham, Dinh S Bui, Caroline J Lodge, Michael J Abramson, Adrian J Lowe, Shuai Li, Aung K Win, Mark Hew, and Shyamali C Dharmage. Genetic ancestry is associated with asthma, and this could be modified by environmental factors. a systematic review. Clinical and Experimental Allergy, 53(6):668, 2023. URL https://onlinelibrary.wiley.com/doi/pdfdirect/10.1111/cea.14308?download=true.

[54] 3rd Washington, C., M. Dapas, A. Biddanda, K. M. Magnaye, I. Aneas, B. A. Helling, B. Szczesny, M. P. Boorgula, M. A. Taub, E. Kenny, R. A. Mathias, K. C. Barnes, G. K. Khurana Hershey, C. M. Kercsmar, J. D. Gereige, M. Makhija, R. S. Gruchalla, M. A. Gill, A. H. Liu, D. Rastogi, W. Busse, P. J. Gergen, C. M. Visness, D. R. Gold, T. Hartert, C. C. Johnson, Jr. Lemanske, R. F., F. D. Martinez, R. L. Miller, D. Ownby, C. M. Seroogy, A. L. Wright, E. M. Zoratti, L. B. Bacharier, M. Kattan, G. T. O’Connor, R. A. Wood, M. A. Nobrega, M. C. Altman, D. J. Jackson, J. E. Gern, C. G. McKennan, and C. Ober. African-specific alleles modify risk for asthma at the 17q12-q21 locus in african americans. Genome Med, 14 (1):112, 2022. ISSN 1756-994x. doi: 10.1186/s13073-022-01114-x. URL https://link.springer.com/content/pdf/10.1186/s13073-022-01114-x.pdf.

[55] Abhijith Biji, Kathleen Ferar, Vikas Pejaver, Eimear E. Kenny, Bian Liu, and Samira Asgari. Integrating social determinants of health and genetic risk in disease risk models. The American Journal of Human Genetics, 113(7):1434–1447, 2026. ISSN 0002-9297. doi: 10.1016/j.ajhg.2026.05.014. URL 10.1016/j.ajhg.2026.05.014.

[56] Arthur E Hoerl and Robert W Kennard. Ridge regression: Biased estimation for nonorthogonal problems. Technometrics, 12(1):55–67, 1970. ISSN 0040-1706.

[57] J. Friedman, T. Hastie, and R. Tibshirani. Regularization paths for generalized linear models via coordinate descent. J Stat Softw, 33(1):1–22, 2010. ISSN 1548-7660 (Print) 1548-7660.

[58] Wen-Hua Wei, Gibran Hemani, and Chris S. Haley. Detecting epistasis in human complex traits. Nature Reviews Genetics, 15(11):722–733, 2014. ISSN 1471-0064. doi: 10.1038/nrg3747. URL 10.1038/nrg3747https://www.nature.com/articles/nrg3747.

[59] Carolyn M. Hutter, Leah E. Mechanic, Nilanjan Chatterjee, Peter Kraft, and Elizabeth M. Gillanders. Gene-environment interactions in cancer epidemiology: A national cancer institute think tank report. Genetic Epidemiology, 37 (7):643–657, 2013. ISSN 0741-0395. doi: 10.1002/gepi.21756. URL https://onlinelibrary.wiley.com/doi/abs/10.1002/gepi.21756https://onlinelibrary.wiley.com/doi/10.1002/gepi.21756.

[60] T. Baltrusaitis, C. Ahuja, and L. P. Morency. Multimodal machine learning: A survey and taxonomy. IEEE Trans Pattern Anal Mach Intell, 41(2):423–443, 2019. ISSN 0098-5589. doi: 10.1109/tpami.2018.2798607.

[61] Ethan Perez, Florian Strub, Harm de Vries, Vincent Dumoulin, and Aaron Courville. Film: visual reasoning with a general conditioning layer, 2018.

[62] Aurélien Beaude, Franck Augé, Farida Zehraoui, and Blaise Hanczar. Crossattomics: multiomics data integration with cross-attention. Bioinformatics, 41(6), 2025. ISSN 1367-4811. doi: 10.1093/bioinformatics/btaf302. URL 10.1093/bioinformatics/btaf302.

[63] Dian Meng, Yu Feng, Kaishen Yuan, Zitong Yu, Qin Cao, Lixin Cheng, and Xubin Zheng. scmmae: masked cross-attention network for single-cell multimodal omics fusion to enhance unimodal omics. Briefings in Bioinformatics, 26(1), 2025. ISSN 1477-4054. doi: 10.1093/bib/bbaf010. URL 10.1093/bib/bbaf010.

[64] Diego Machado Reyes, Myson Burch, Laxmi Parida, and Aritra Bose. A foundation model for learning genetic associations from brain imaging phenotypes. Bioinformatics Advances, 5(1), 2025. ISSN 2635-0041. doi: 10.1093/bioadv/vbaf196. URL 10.1093/bioadv/vbaf196.

[65] Jinghui Liu, Daniel Capurro, Anthony Nguyen, and Karin Verspoor. Attention-based multimodal fusion with contrast for robust clinical prediction in the face of missing modalities. Journal of Biomedical Informatics, 145:104466, 2023. ISSN 1532-0464. doi: 10.1016/j.jbi.2023.104466. URL https://www.sciencedirect.com/science/article/pii/S1532046423001879.

[66] Yao-Hung Hubert Tsai, Shaojie Bai, Paul Pu Liang, J Zico Kolter, Louis-Philippe Morency, and Ruslan Salakhutdinov. Multimodal transformer for unaligned multimodal language sequences. In Proceedings of the 57th annual meeting of the association for computational linguistics, pages 6558–6569.

[67] Fengmao Lv, Xiang Chen, Yanyong Huang, Lixin Duan, and Guosheng Lin. Progressive modality reinforcement for human multimodal emotion recognition from unaligned multimodal sequences. In Proceedings of the IEEE/CVF conference on computer vision and pattern recognition, pages 2554–2562.

[68] Wei Han, Hui Chen, Alexander Gelbukh, Amir Zadeh, Louis-philippe Morency, and Soujanya Poria. Bi-bimodal modality fusion for correlation-controlled multimodal sentiment analysis. In Proceedings of the 2021 international conference on multimodal interaction, pages 6–15.

[69] Sarthak Jain and Byron C. Wallace. Attention is not explanation. Proceedings of the 2019 Conference of the North American Chapter of the Association for Computational Linguistics: Human Language Technologies, Volume 1 (Long and Short Papers), pages 3543–3556. Association for Computational Linguistics. doi: 10.18653/v1/N19-1357. URL https://aclanthology.org/N19-1357/https://doi.org/10.18653/v1/N19-1357https://aclanthology.org/N19-1357.pdf.

[70] Sofia Serrano and Noah A. Smith. Is attention interpretable? Proceedings of the 57th Annual Meeting of the Association for Computational Linguistics, pages 2931–2951. Association for Computational Linguistics. doi: 10.18653/v1/P19-1282. URL https://aclanthology.org/P19-1282/https://doi.org/10.18653/v1/P19-1282https://aclanthology.org/P19-1282.pdf.

[71] Sarah Wiegreffe and Yuval Pinter. Attention is not not explanation. Proceedings of the 2019 Conference on Empirical Methods in Natural Language Processing and the 9th International Joint Conference on Natural Language Processing (EMNLP-IJCNLP), pages 11–20. Association for Computational Linguistics. doi: 10.18653/v1/D19-1002. URL https://aclanthology.org/D19-1002/https://doi.org/10.18653/v1/D19-1002https://aclanthology.org/D19-1002.pdf.

[72] Gherman Novakovsky, Nick Dexter, Maxwell W. Libbrecht, Wyeth W. Wasserman, and Sara Mostafavi. Obtaining genetics insights from deep learning via explainable artificial intelligence. Nature Reviews Genetics, 24(2): 125–137, 2023. ISSN 1471-0064. doi: 10.1038/s41576-022-00532-2. URL 10.1038/s41576-022-00532-2 https://www.nature.com/articles/s41576-022-00532-2.

[73] Haitham A. Elmarakeby, Justin Hwang, Rand Arafeh, Jett Crowdis, Sydney Gang, David Liu, Saud H. AlDubayan, Keyan Salari, Steven Kregel, Camden Richter, Taylor E. Arnoff, Jihye Park, William C. Hahn, and Eliezer M. Van Allen. Biologically informed deep neural network for prostate cancer discovery. Nature, 598(7880):348–352, 2021. ISSN 1476-4687. doi: 10.1038/s41586-021-03922-4. URL 10.1038/s41586-021-03922-4 https://www.nature.com/articles/s41586-021-03922-4.pdf.

[74] Mukund Sundararajan, Ankur Taly, and Qiqi Yan. Axiomatic attribution for deep networks, 2017.

[75] Scott M. Lundberg and Su-In Lee. A unified approach to interpreting model predictions, 2017.

[76] Jason Yosinski, Jeff Clune, Yoshua Bengio, and Hod Lipson. How transferable are features in deep neural networks?, 2014.

[77] Ali Sharif Razavian, Hossein Azizpour, Josephine Sullivan, and Stefan Carlsson. Cnn features off-the-shelf: An astounding baseline for recognition, 2014. URL 10.1109/CVPRW.2014.131.

[78] Yunchuan Kong and Tianwei Yu. A deep neural network model using random forest to extract feature representation for gene expression data classification. Scientific Reports, 8(1):16477, 2018. ISSN 2045-2322. doi: 10.1038/s41598-018-34833-6. URL 10.1038/s41598-018-34833-6 https://www.nature.com/articles/s41598-018-34833-6.pdf.

[79] Riccardo Miotto, Li Li, Brian A. Kidd, and Joel T. Dudley. Deep patient: An unsupervised representation to predict the future of patients from the electronic health records. Scientific Reports, 6(1):26094, 2016. ISSN 2045-2322. doi: 10.1038/srep26094. URL 10.1038/srep26094https://www.nature.com/articles/srep26094.pdf.

[80] T. A. Suleiman, D. T. Anyimadu, A. D. Permana, H. A. A. Ngim, and A. Scotto di Freca. Two-step hierarchical binary classification of cancerous skin lesions using transfer learning and the random forest algorithm. Vis Comput Ind Biomed Art, 7(1): 15, 2024. ISSN 2096-496X (Print) 2524-4442. doi: 10.1186/s42492-024-00166-7. URL https://link.springer.com/content/pdf/10.1186/s42492-024-00166-7.pdf.

[81] R. Jose, R. Rooney, N. Nagisetty, R. Davis, and D. Hains. Biorepository and integrative genomics initiative: designing and implementing a preliminary platform for predictive, preventive and personalized medicine at a pediatric hospital in a historically disadvantaged community in the usa. Epma j, 9(3):225–234, 2018. ISSN 1878-5077 (Print) 1878-5077. doi: 10.1007/s13167-018-0141-y. URL https://link.springer.com/article/10.1007/s13167-018-0141-y.

[82] Dinh S Bui, John A Burgess, Adrian J Lowe, Jennifer L Perret, Caroline J Lodge, Minh Bui, Stephen Morrison, Bruce R Thompson, Paul S Thomas, and Graham G Giles. Childhood lung function predicts adult chronic obstructive pulmonary disease and asthma–chronic obstructive pulmonary disease overlap syndrome. American journal of respiratory and critical care medicine, 196(1):39–46, 2017. ISSN 1073-449X.

[83] Global Initiative for Asthma. Global strategy for asthma management and preventionglobal initiative for asthma. Report, 2024. URL https://ginasthma.org/wp-content/uploads/2024/12/GINA-Summary-Guide-2024-WEB-WMS.pdf.

[84] Robin J. Hofmeister, Diogo M. Ribeiro, Simone Rubinacci, and Olivier Delaneau. Accurate rare variant phasing of whole-genome and whole-exome sequencing data in the uk biobank. Nature Genetics, 55(7):1243–1249, 2023. ISSN 1546-1718. doi: 10.1038/s41588-023-01415-w. URL 10.1038/s41588-023-01415-w.

[85] Adam Auton, Gonçalo R. Abecasis, David M. Altshuler, Richard M. Durbin, Gonçalo R. Abecasis, David R. Bentley, Aravinda Chakravarti, Andrew G. Clark, Peter Donnelly, Evan E. Eichler, Paul Flicek, Stacey B. Gabriel, Richard A. Gibbs, Eric D. Green, Matthew E. Hurles, Bartha M. Knoppers, Jan O. Korbel, Eric S. Lander, Charles Lee, Hans Lehrach, Elaine R. Mardis, Gabor T. Marth, Gil A. McVean, Deborah A. Nickerson, Jeanette P. Schmidt, Stephen T. Sherry, Jun Wang, Richard K. Wilson, Richard A. Gibbs, Eric Boerwinkle, Harsha Doddapaneni, Yi Han, Viktoriya Korchina, Christie Kovar, Sandra Lee, Donna Muzny, Jeffrey G. Reid, Yiming Zhu, Jun Wang, Yuqi Chang, Qiang Feng, Xiaodong Fang, Xiaosen Guo, Min Jian, Hui Jiang, Xin Jin, Tianming Lan, Guoqing Li, Jingxiang Li, Yingrui Li, Shengmao Liu, Xiao Liu, Yao Lu, Xuedi Ma, Meifang Tang, Bo Wang, Guangbiao Wang, Honglong Wu, Renhua Wu, Xun Xu, Ye Yin, Dandan Zhang, Wenwei Zhang, Jiao Zhao, Meiru Zhao, Xiaole Zheng, Eric S. Lander, David M. Altshuler, Stacey B. Gabriel, Namrata Gupta, Neda Gharani, Lorraine H. Toji, Norman P. Gerry, Alissa M. Resch, Paul Flicek, Jonathan Barker, Laura Clarke, Laurent Gil, Sarah E. Hunt, Gavin Kelman, Eugene Kulesha, Rasko Leinonen, William M. McLaren, Rajesh Radhakrishnan, Asier Roa, Dmitriy Smirnov, Richard E. Smith, Ian Streeter, Anja Thormann, Iliana Toneva, Brendan Vaughan, Xiangqun Zheng-Bradley, David R. Bentley, Russell Grocock, Sean Humphray, Terena James, Zoya Kingsbury, Hans Lehrach, Ralf Sudbrak, Marcus W. Albrecht, et al. A global reference for human genetic variation. Nature, 526(7571):68–74, 2015. ISSN 1476-4687. doi: 10.1038/nature15393. URL 10.1038/nature15393.

[86] Shaun Purcell, Benjamin Neale, Kathe Todd-Brown, Lori Thomas, Manuel AR Ferreira, David Bender, Julian Maller, Pamela Sklar, Paul IW De Bakker, and Mark J Daly. Plink: a tool set for whole-genome association and population-based linkage analyses. The American journal of human genetics, 81(3):559–575, 2007. ISSN 0002-9297.

[87] Shaun Purcell. Plink, 2024. URL http://pngu.mgh.harvard.edu/purcell/plink/.

[88] Sofia Mongardi, Silvia Cascianelli, and Marco Masseroli. Biologically weighted lasso: enhancing functional interpretability in gene expression data analysis. Bioinformatics, 40(10):btae605, 2024. ISSN 1367-4811.

[89] Anna M Ehlers, Idaira M Guerrero-Fonseca, Christophe Altier, Bryan G Yipp, and Sebastien Talbot. Wired for immunity: neuroimmune control of the lung by sensory neurons. Nature Reviews Neuroscience, pages 1–13, 2026. ISSN 1471-003X.

[90] Michael Ashburner, Catherine A Ball, Judith A Blake, David Botstein, Heather Butler, J Michael Cherry, Allan P Davis, Kara Dolinski, Selina S Dwight, and Janan T Eppig. Gene ontology: tool for the unification of biology. Nature genetics, 25(1):25–29, 2000. ISSN 1546-1718. URL https://www.nature.com/articles/ng0500_25.

[91] Suzi A Aleksander, James Balhoff, Seth Carbon, J Michael Cherry, Harold J Drabkin, Dustin Ebert, Marc Feuermann, Pascale Gaudet, and Nomi L Harris. The gene ontology knowledgebase in 2023. Genetics, 224(1):iyad031, 2023. ISSN 1943-2631.

[92] M. Kanehisa and S. Goto. Kegg: kyoto encyclopedia of genes and genomes. Nucleic Acids Res, 28(1):27–30, 2000. ISSN 0305-1048 (Print) 0305-1048. doi: 10.1093/nar28.1.27.

[93] D. J. Rigden and X. M. Fernández. The 2024 nucleic acids research database issue and the online molecular biology database collection. Nucleic Acids Res, 52(D1): D1–d9, 2024. ISSN 0305-1048 (Print) 0305-1048. doi: 10.1093/nar/gkad1173.

[94] Edward Y Chen, Christopher M Tan, Yan Kou, Qiaonan Duan, Zichen Wang, Gabriela Vaz Meirelles, Neil R Clark, and Avi Ma’ayan. Enrichr: interactive and collaborative html5 gene list enrichment analysis tool. BMC bioinformatics, 14(1): 128, 2013. ISSN 1471-2105.

[95] Maxim V Kuleshov, Matthew R Jones, Andrew D Rouillard, Nicolas F Fernandez, Qiaonan Duan, Zichen Wang, Simon Koplev, Sherry L Jenkins, Kathleen M Jagodnik, and Alexander Lachmann. Enrichr: a comprehensive gene set enrichment analysis web server 2016 update. Nucleic acids research, 44(W1):W90–W97, 2016. ISSN 0305-1048.

[96] Antonella Zanobetti, Patrick H Ryan, Brent A Coull, Heike Luttmann-Gibson, Soma Datta, Jeffrey Blossom, Cole Brokamp, Nathan Lothrop, Rachel L Miller, and Paloma I Beamer. Early-life exposure to air pollution and childhood asthma cumulative incidence in the echo crew consortium. JAMA Network Open, 7(2): e240535, 2024. ISSN 2574-3805.

[97] Jocelyn R Grunwell, Cydney Opolka, Carrie Mason, and Anne M Fitzpatrick. Geospatial analysis of social determinants of health identifies neighborhood hot spots associated with pediatric intensive care use for life-threatening asthma. The Journal of Allergy and Clinical Immunology: In Practice, 10(4):981–991. e1, 2022. ISSN 2213-2198.

[98] Adolfo L Molina, Yamilé Molina, Susan C Walley, Chang L Wu, Aowen Zhu, and Gabriela R Oates. Residential instability, neighborhood deprivation, and pediatric asthma outcomes. Pediatric pulmonology, 55(6):1340–1348, 2020. ISSN 8755-6863. URL https://onlinelibrary.wiley.com/doi/10.1002/ppul.24771.

[99] Joshua A Lawson, Donna C Rennie, Don W Cockcroft, Roland Dyck, Anna Afanasieva, Oluwafemi Oluwole, and Jinnat Afsana. Childhood asthma, asthma severity indicators, and related conditions along an urban-rural gradient: a cross-sectional study. BMC pulmonary medicine, 17(1):4, 2017. ISSN 1471-2466. URL https://link.springer.com/content/pdf/10.1186/s12890-016-0355-5.pdf.

[100] Esteban Correa-Agudelo, Lili Ding, Andrew F Beck, Cole Brokamp, Mekibib Altaye, Robert S Kahn, and Tesfaye B Mersha. Understanding racial disparities in childhood asthma using individual-and neighborhood-level risk factors. Journal of Allergy and Clinical Immunology, 150(6):1427–1436. e5, 2022. ISSN 0091-6749.

[101] Matthew Tancik, Pratul Srinivasan, Ben Mildenhall, Sara Fridovich-Keil, Nithin Raghavan, Utkarsh Singhal, Ravi Ramamoorthi, Jonathan Barron, and Ren Ng. Fourier features let networks learn high frequency functions in low dimensional domains. Advances in neural information processing systems, 33:7537–7547, 2020.

[102] Lukas Biewald. Experiment tracking with weights and biases, 2020. URL https://wandb.com.

[103] Narine Kokhlikyan, Vivek Miglani, Miguel Martin, Edward Wang, Bilal Alsallakh, Jonathan Reynolds, Alexander Melnikov, Natalia Kliushkina, Carlos Araya, and Siqi Yan. Captum: A unified and generic model interpretability library for pytorch. arXiv preprint arXiv:2009.07896, 2020.

[104] Nicolai Meinshausen and Peter Bühlmann. Stability selection. Journal of the Royal Statistical Society Series B: Statistical Methodology, 72(4):417–473, 2010. ISSN 1369-7412.

[105] Milton Pividori, Nathan Schoettler, Dan L Nicolae, Carole Ober, and Hae Kyung Im. Shared and distinct genetic risk factors for childhood-onset and adult-onset asthma: genome-wide and transcriptome-wide studies. The Lancet Respiratory Medicine, 7 (6):509–522, 2019. ISSN 2213-2600.

[106] Pinja Ilmarinen, Leena E Tuomisto, and Hannu Kankaanranta. Phenotypes, risk factors, and mechanisms of adult-onset asthma. Mediators of inflammation, 2015(1): 514868, 2015. ISSN 1466-1861.

