## Supplementary Information for "X-Admix: An Interpretable Multimodal Cross-Attention Framework for Integrating Genotype, Local Ancestry, and Social Drivers of Health in Admixed African American Populations"

### Supplementary Material (File S1)

Table S1: Per-fold feature pool sizes for each data modality.

**(a) BIG Cohort**

| Fold | Genotype | LA | SDOH |
|---|---|---|---|
| 1 | 13 | 264 | 4 |
| 2 | 17 | 208 | 4 |
| 3 | 15 | 238 | 4 |
| 4 | 15 | 241 | 4 |
| 5 | 12 | 233 | 4 |
| 6 | 12 | 245 | 4 |
| 7 | 11 | 230 | 4 |
| 8 | 11 | 253 | 4 |
| 9 | 18 | 235 | 4 |
| 10 | 13 | 264 | 4 |

**(b) AoU Cohort**

| Fold | Genotype | LA | SDOH |
|---|---|---|---|
| 1 | 15 | 16 | 5 |
| 2 | 27 | 18 | 5 |
| 3 | 58 | 39 | 5 |
| 4 | 36 | 34 | 5 |
| 5 | 50 | 22 | 5 |
| 6 | 79 | 27 | 5 |
| 7 | 33 | 11 | 5 |
| 8 | 39 | 36 | 5 |
| 9 | 13 | 24 | 5 |
| 10 | 57 | 25 | 5 |

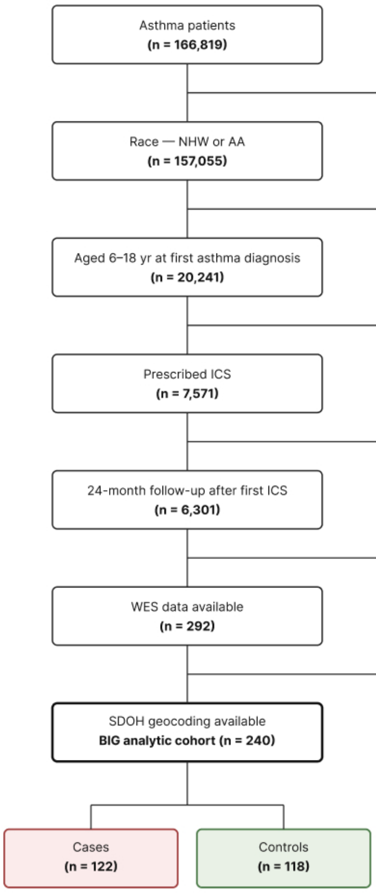


Figure S1: Consort diagram for the BIG pediatric cohort, resulting in a cohort of 240 participants.

Table S2: Performance of different model configurations on the BIG cohort. Values are reported as mean ± SD across folds.

| Model Configuration | ROC AUC | Precision | Recall |
|---|---|---|---|
| $S_{\text{Geno}\leftarrow\text{SDOH}}$ | 0.748 ± 0.173 | 0.680 ± 0.143 | 0.709 ± 0.216 |
| $S_{\text{LA}\leftarrow\text{SDOH}}$ | 0.606 ± 0.117 | 0.579 ± 0.092 | 0.588 ± 0.160 |
| $S_{\text{Geno}\leftarrow\text{LA}}$ | 0.639 ± 0.127 | 0.623 ± 0.121 | 0.633 ± 0.150 |

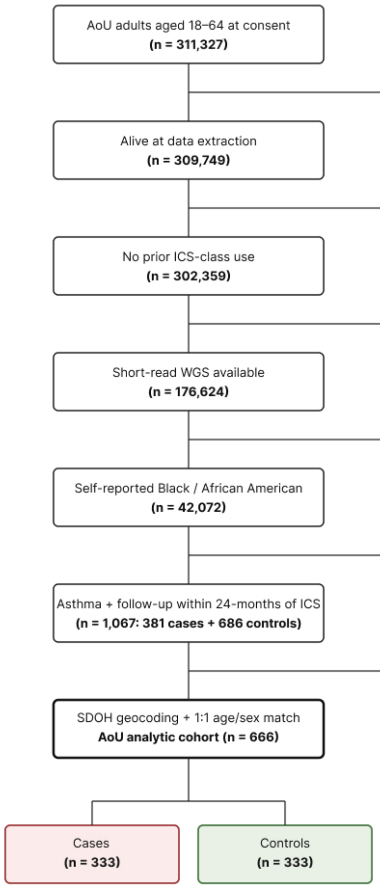


Figure S2: Consort diagram for the All of Us (AoU) adult replication cohort, resulting in 666 participants.

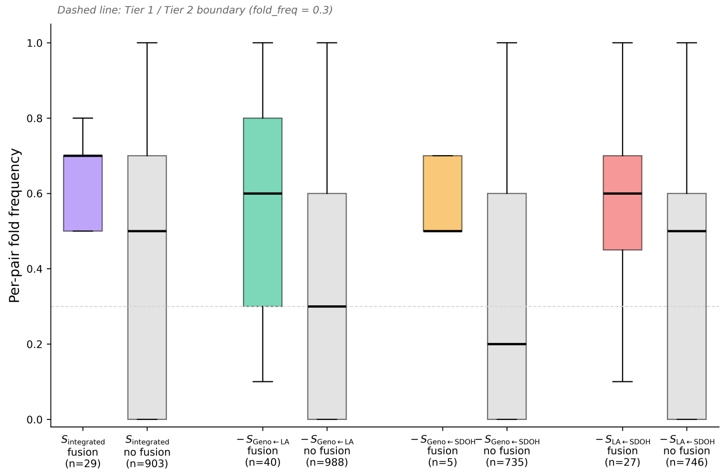


Figure S3: Per-pair fold-frequency distributions across model configurations. Distributions are shown for $S_{\text{integrated}}$ and the three single-stream ablations ($-S_{\text{Geno}\leftarrow\text{LA}}$, $-S_{\text{Geno}\leftarrow\text{SDOH}}$, $-S_{\text{LA}\leftarrow\text{SDOH}}$), stratified by whether each pair receive Grad × Activation (fusion-side attribution) support (coloured) or is identified by SHAP (RF attribution) alone (grey). Fold-frequency is the fraction of the 10 outer cross-validation folds in which a pair appears in the consensus pool. The dashed horizontal line at *fold_freq* = 0.3 marks the Tier 1 / Tier 2 boundary. Sample sizes are listed below each box. The right-shift of fusion-supported pairs relative to fusion-negative pairs across all four configurations confirms that fusion attribution preferentially recovers fold-stable consensus pairs.

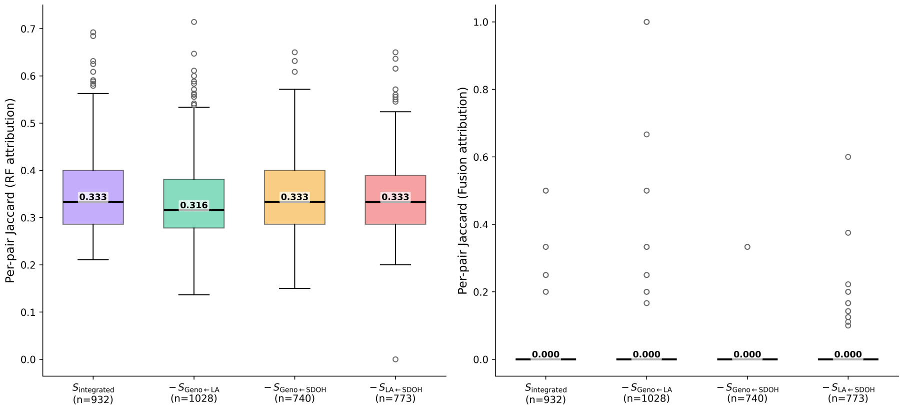


Figure S4: Per-pair Jaccard similarity distributions across model configurations. Distributions are shown separately for RF attribution (left) and fusion attribution (right). Each box summarises the within-fold Jaccard overlap of the two features in a pair across the $z$-dimensions in which they co-occur, computed over each ablation's own retained consensus pairs (n); annotated values give the median for each configuration. RF Jaccard distributions are graded, with medians clustered tightly at 0.316 ($-S_{\text{Geno}\leftarrow\text{LA}}$) to 0.333 ($S_{\text{integrated}}$, $-S_{\text{Geno}\leftarrow\text{SDOH}}$, $-S_{\text{LA}\leftarrow\text{SDOH}}$). Fusion Jaccard distributions collapse to a median of 0.000 across all four configurations, with non-zero values appearing only as sparse outliers. The asymmetry between the two panels reflects the difference in candidate pool structure between the two channels: Grad × Activation assigns each dimension its own 10-feature list, whereas SHAP draws every dimension from a shared 120-feature pool.

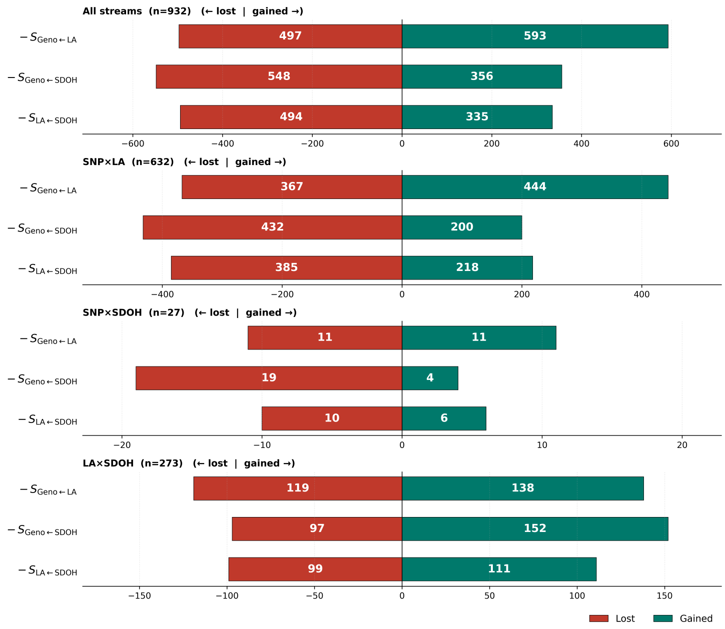


Figure S5: Cross-modal pair turnover under single-stream ablation, total and stratified by interaction type. For each of the three single-stream ablations ($-S_{Geno\leftarrow LA}$, $-S_{Geno\leftarrow SDOH}$, $-S_{LA\leftarrow SDOH}$), bars show pairs lost from the $S_{integrated}$ consensus pool (red, negative axis) and compensatory pairs gained (green, positive axis). Top panel: total turnover over all 932 $S_{integrated}$ pairs. Removing $S_{Geno\leftarrow LA}$ is the only ablation under which gain exceeds loss (593 vs. 497; net +96, +10.3%); removing $S_{Geno\leftarrow SDOH}$ produces the largest absolute loss (548) and the largest net contraction (net −192, −20.6%); removing $S_{LA\leftarrow SDOH}$ produces an intermediate net contraction (net −159, −17.1%). Lower panels: turnover stratified by interaction type. Geno ← LA ($n = 632$ in $S_{integrated}$): loses most heavily under $-S_{Geno\leftarrow SDOH}$ (432 lost; net −232), indicating that the Geno ← LA pair catalogue is anchored not only by the dedicated $S_{Geno\leftarrow LA}$ stream but also by routing through $S_{Geno\leftarrow SDOH}$. LA ← SDOH ($n = 273$): grows under every ablation (net +19 to +55), with the largest expansion under $-S_{Geno\leftarrow SDOH}$ (152 gained vs. 97 lost), consistent with environmental signal being rerouted through the ancestry–SDOH axis when the direct genotype–SDOH route is unavailable. Geno ← SDOH ($n = 27$): quantitatively negligible across all ablations and shown for completeness.
